# Integrative analysis of pre-treatment RNA expression signatures and recurrent genomic alterations: Link to menopausal status, short-term endocrine therapy response and disease-free survival in luminal breast cancer

**DOI:** 10.1101/2025.06.11.25329354

**Authors:** Guokun Zhang, Hua Ni, Lidiya Mishieva, Stephan Bartels, Matthias Christgen, Henriette Christgen, Leonie Donata Kandt, Mieke Raap, Ronald E. Kates, Oleg Gluz, Monika Graeser, Sherko Kümmel, Ulrike Nitz, Christoph Plass, Ulrich Mansmann, Christine zu Eulenburg, Clarissa Gerhäuser, Hans H. Kreipe, Nadia Harbeck

**Author notes:** Corresponding author: Dr. Clarissa Gerhäuser, Division of Cancer Epigenomics, German Cancer Research Center, Im Neuenheimer Feld 280, 69120 Heidelberg, Germany. HN: Department of Pathology, School of Basic Medical Sciences, Peking University Third Hospital, Peking University Health Science Center. Social media description: P53 mutations, genomic & immune signatures predict therapy resistance & long-term outcomes in luminal breast cancer.

## Abstract

**Background:** Endocrine therapy with tamoxifen (TAM) or aromatase inhibitors (AI) is an effective treatment for patients with estrogen receptor-positive, HER2-negative luminal breast cancer. However, many patients do not respond to this therapy, leading to disease recurrence. This study aimed to identify baseline clinical, molecular, and genetic features associated with menopause status, primary endocrine therapy resistance and long-term outcomes in luminal breast cancer.

**Methods:** We analyzed 220 patients from the WSG-ADAPT trial with early-stage, estrogen receptor-positive, HER2-negative breast cancer, who received three weeks of preoperative endocrine therapy with TAM or AI. Pre-treatment tumor samples were profiled using the NanoString BC360 panel, and post-treatment samples were analyzed for recurrent genomic alterations by next generation panel sequencing. A subset of the TCGA-BRCA cohort was used for external validation. Univariate Cox regression analyses were used for prognosis analysis.

**Results:** The NanoString signatures were clustered into three stable blocks: A (reactive microenvironment and stemness), B (immune) and C (proliferation and genomic risk). Non-responders more frequently harbored *TP53* mutations, which were linked to significantly elevated pro-tumorigenic immune-(IFN-γ, Inflammatory Chemokines, Macrophages and Treg) and proliferation-related (BC Proliferation, Genomic Risk, and homologous recombination deficiency (HRD)) signature scores. In the AI group, signatures associated with reduced disease-free survival included BC p53 (hazard ratio [HR] with 95% confidence interval [CI] = 2.74 [1.08-6.94]), Genomic Risk (HR = 2.5 [1.07-5.83]), HRD (HR = 2.44 [1.12-5.29]) and Hypoxia (HR = 2.12 [1.17-3.87]). High expression of PD-1 (HR = 0.44 [0.21-0.94]) and PGR (HR = 0.24 [0.07-0.81]) indicated better outcomes, respectively. These associations were validated using external data.

**Conclusions:** Endocrine resistance in luminal breast cancer is characterized by elevated immune signatures, increased proliferation, and specific genomic alterations. The integration of clinical information, gene expression patterns, and genetic data enhances patient stratification and potentially informs treatment decisions. These findings support the use of integrative analyses to guide personalized endocrine therapy and improve outcomes.

**Trial registration:** WSG-ADAPT, NCT01779206, Registered 2013-01-25, retrospectively registered.

**Highlights:**

- Mutations in tumor protein p53 linked to endocrine therapy resistance in luminal breast cancer.
- Non-responders have higher pre-treatment immune activity and cell proliferation signals in tumors.
- Poor survival is associated with signatures of mutant p53, genomic risk, repair deficiency and hypoxia in tumors.
- High levels of programmed cell death protein 1 and progesterone receptor predict better outcomes.
- Combining clinical, transcriptomic, and genetic data may improve personalized therapy decisions.

## Background

Estrogen receptor-positive (ER+) breast cancer accounts for approximately 75% of breast cancer diagnoses and relies primarily on estrogen signaling for tumor growth and progression.^1^ Standard treatments for ER+ breast cancer include endocrine therapies such as selective estrogen receptor modulators (SERMs) like tamoxifen (TAM) and aromatase inhibitors (AI), which block estrogen production.^2^ Endocrine resistance is a significant clinical challenge because up to 40% of patients experience disease relapse during or after endocrine therapy.^3^ Thus, understanding the mechanisms underlying endocrine resistance is essential for improving patient prognosis and informing treatment decisions.

Endocrine resistance in ER+ breast cancer is broadly categorized as primary resistance, where tumors fail to respond to initial hormone therapy, or secondary resistance, which emerges after an initial period of responsiveness.^4,5^ This study focuses on primary resistance, which is determined by tumor-intrinsic and microenvironmental factors present before the initiation of endocrine therapy.

The West German Study Group Adjuvant Dynamic Marker-Adjusted Personalized Therapy (WSG-ADAPT) trial has enrolled more than 5,600 patients with luminal breast cancer (BC) to date. This trial provides a novel approach to investigating endocrine resistance and includes 2,290 patients with early-stage, ER-positive, HER2-negative breast cancer.^6^ Patients receive a short, three-week course of preoperative endocrine therapy (pET) with TAM in pre- and AI in postmenopausal patients. Treatment response is assessed using Ki67 immunohistochemistry as a marker of tumor proliferation before and after pET. Patients are classified as responders or non-responders based on changes in Ki67 levels. This design offers a unique opportunity to investigate pre-treatment baseline tumor biology and gene expression in relation to short-term endocrine response and long-term outcome.

Accumulating evidence highlights the value of integrative transcriptomic approaches for identifying biological pathways associated with primary endocrine resistance. Together, these studies suggest that primary endocrine resistance is associated with molecular features and distinct gene expression programs, including aberrant estrogen signaling,^7^ a hypoxic tumor microenvironment,^8^ and immune activation.^8^ These findings support the development of integrated biomarker strategies to inform early treatment decisions in ER+ breast cancer.

The NanoString Breast Cancer 360™ (BC360) panel analyzes the expression of 758 curated genes relevant to breast cancer biology. These genes are stratified into 42 NanoString signatures that reflect tumor-intrinsic processes, immune activity, tumor microenvironment composition, and key breast cancer pathways.^9^ The signature panel provides an opportunity to explore prognostic and predictive markers of endocrine responsiveness.

The Oncotype DX Recurrence Score (RS) estimates the risk of distant recurrence based on the mRNA expression of 21 genes^10^ and is widely used in the clinics to guide treatment decisions, particularly regarding the need for adjuvant chemotherapy in ER+ breast cancer patients.^11^ Based on the RS, patients are categorized into low-, intermediate-, and high-risk groups, enabling a more individualized treatment approach.

Our study aimed to identify molecular signatures associated with endocrine resistance in pre- and postmenopausal patients and to uncover potential mechanisms by integrating comprehensive transcriptomic profiling from the BC360 panel with information on therapy response, RS groupings, and clinical and genomic data. Our findings may inform personalized therapy strategies for ER+ breast cancer.

## Methods

### NanoString cohort

We analyzed 220 patients (NanoString cohort, Supplementary Table S1) with ER+ and/or PR+, HER2− tumors enrolled in the phase II WSG-ADAPT trial (NCT01779206). Responders (R) to preoperative endocrine therapy (pET) were defined as post-pET Ki67 < 10% and a ≥ 70% reduction from baseline; non-responders (NR) had post-pET Ki67 ≥ 20% and ≤ 20% reduction. R and NR were initially matched for baseline histopathological features as described previously.^12^ Pairing was partially broken due to RNA availability (Supplementary Table S2). Stromal tumor-infiltrating lymphocytes (TILs) were scored on baseline formalin-fixed, paraffin-embedded (FFPE) biopsies and categorized into three groups: 0-9%, 10-40%, and 41-100%.

### TCGA-BRCA subcohort

A subset (n=260, Supplementary Tables S2-S3) of TCGA-BRCA^13,14^ cohort was used as a validation cohort, subsampled to match the NanoString cohort as reported.^12^ Basal-like samples were excluded. Missing race annotations were inferred from DNA methylation data (R package SeSAMe v1.10.4).

### NanoString gene expression profiling

RNA was extracted from diagnostic FFPE biopsies. Gene expression was quantified using the NanoString Breast Cancer 360™ (BC360) panel (Supplementary Table S2, gene list in Ref.^15^). Quality control and normalization were performed by NanoString using nSolver 4.0 and an in-house pipeline; samples with irregular control probe distributions were excluded. PAM50 molecular subtypes were assigned by centroid correlation^16^. Genomic Risk scores were computed from PAM50 and proliferation signatures using genomic-only methods and scaled 0-100.^9^ For TCGA-BRCA, signature scores were calculated as the mean log2 expression of genes in each NanoString signature.

### Correlation of NanoString signature scores

Spearman correlations were calculated between BC360 signature scores, clinico-pathological features, Recurrence Score (RS) groups, NanoString PAM50 correlations, and the Predictive Endocrine Resistance Index (PERCI),^12^ a composite score integrating clinico-pathological, tumour microenvironment composition, genomic, and epigenomic variables to capture primary endocrine resistance. In the TCGA-BRCA subcohort, correlations were restricted to signature scores and PERCI-450k. Hierarchical clustering (ward.D2 linkage) was used to group correlated features. Correlation differences between treatment groups were tested (R package diffcor v0.8.4).^17^

### Mutation profiling

Post-pET tumors were analyzed by targeted next-generation sequencing as described.^12^ We prioritized the 25 most recurrent genomic alterations (RGAs) with a frequency ≥ 7.5% in the NanoString treatment-response groups. RGA frequencies were compared between treatment-response and menopausal groups. Differential expression between wild-type and altered groups was tested using the Wilcoxon rank sum.

### Differential signature score analysis

Linear mixed models were used to compare signature scores between treatment-response, RS risk and menopausal groups, including patient pair ID as a random effect where applicable (R package variancePartition v1.38.0). Comparisons were adjusted for clinico-pathological covariates that differed significantly between groups. Results are reported as log2 fold changes with 95% confidence intervals and adjusted *p*-values.

### Partial Least Squares Discriminant Analysis (PLS-DA)

PLS-DA models to discriminate between treatment-response groups were fitted on baseline BC360 signature scores, RGAs, and age (R package mixOmics v6.32.0).^18,19^ Qualitative variables were dummy-coded; levels with < 5% prevalence and quantitative variables with < 5% mean absolute deviation were excluded. Model performance was assessed by AUC of the ROC curve (R package pROC v1.19.0.1). Sparse PLS-DA with lasso penalization was applied for feature selection, with stable features identified using 50x repeated cross-validation. The top 20 features were identified by selection frequency.

### Survival analysis

Associations between baseline BC360 signatures and survival outcomes were assessed using Cox proportional hazards models on z-transformed scores. Results are presented as hazard ratios with 95% confidence intervals. No multiple testing correction was applied. Associations were validated using KM Plotter.^20^

### Statistical analysis

Significant associations between categorical variables were assessed using Fisher’s exact test unless otherwise specified. Clinico-pathological characteristics between responder groups were evaluated using a Cumulative Link Mixed Model for ordinal variables (R package ordinal v2023.12-4.1) and a Generalized Linear Mixed-Effects Model for binary variables (R package lme4 v1.1-37), with patient pair ID as a random effect where applicable. Comparisons of Ki67 at baseline between treatment groups were conducted using chi-squared tests. Variance of log2-transformed signature scores across menopausal, treatment, and response groups was assessed and visualized. Unless otherwise noted, *p*-values were adjusted for multiple testing to control the false discovery rate (FDR) using the Benjamini-Hochberg method. Plots were generated with ggplot2 (v4.0.0) unless stated otherwise. RGAs were summarized with OncoPrints, and differential signature expression between wild-type and altered groups was visualized with heatmaps (R package ComplexHeatmap v2.24.1). Significant differential signatures and survival hazard ratios were displayed as forest plots (R package forestplot v3.1.7). Overall analyses are exploratory and hypothesis-generating.

## Results

### Clinico-pathological and molecular characteristics of the cohorts

We analyzed a cohort of 220 patients from the WSG-ADAPT trial, who were stratified by menopause status and prospective pET response into TAM and AI responders (R) and non-responders (NR). A comparison of clinico-pathological features revealed major differences between the treatment groups, besides age as a proxy of menopausal status. Cases in the AI group had significantly higher histological grades and Ki67 staining and were more likely to be of the luminal B subtype or in the high-risk group RS3. Infiltrating lobular breast cancer (ILBC) subtype and high PR staining were more prevalent in the premenopausal TAM group (Supplementary Fig. S1). Nonetheless, clinico-pathological features between the response groups were well balanced, supporting the comparability of the response groups. In the AI group, non-responders were significantly younger than responders (*p* = 0.004). PAM50 subtyping revealed a significant enrichment of the Luminal B subtype in TAM non-responders, while tumor-infiltrating lymphocytes (TILs) were significantly lower in TAM non-responders.

In the TCGA-BRCA subcohort selected as a validation cohort, the cases did not show significant differences in clinico-pathological characteristics between menopausal groups, except for age, stage and race. Compared to the NanoString cohort, this cohort had more cases of ILBC and Luminal A subtype. Overall, the cases were at higher histological stage, but had lower grade, and about 30% had equivocal HER2 staining (Supplementary Table S4).

### Reduced PGR and elevated ESR1 expression in postmenopausal cases

The genes covered by the NanoString BC360 array are partly assigned to 42 transcriptional signatures (overview in Supplementary Fig. S2 and S3, Supplementary Table S2). We observed greater variability in signature scores in the AI group than the TAM group. In the TCGA-BRCA subcohort, signature scores were overall more variable (Supplementary Fig. S4). In both cohorts, the progesterone receptor (PGR) exhibited the greatest variability and the greatest expression loss in postmenopausal cases (log2FC: NanoString −0.47, TCGA-BRCA −0.72, Supplementary Fig. S5, Supplementary Table S2). Conversely, ESR1 scores were significantly higher in postmenopausal cases (log2FC: NanoString 0.72, TCGA-BRCA 1.22).

### Baseline NanoString signature scores correlate with key clinical and molecular features

To investigate the interactions between baseline signatures and clinico-pathological characteristics, we performed Spearman’s correlation analyses (Supplementary Table S2). Unsupervised clustering grouped the signatures into three functional blocks (A, B, and C) with high positive correlations within the blocks (Fig. 1). Clustering and block annotations were largely confirmed in the TCGA-BRCA subcohort (Supplementary Fig. S6). Additional signatures that did not form strong clusters were grouped into Blocks D (“Tumor-related”) and Block E (“Hormone- and breast cancer-related”) according to NanoString’s predefined biological categories (Supplementary Table S2). There was excellent concordance between percent positive staining of progesterone receptor (PR) at baseline and PGR signature scores (TAM: ρ = 0.6; AI: ρ = 0.79).

**Fig. 1:**
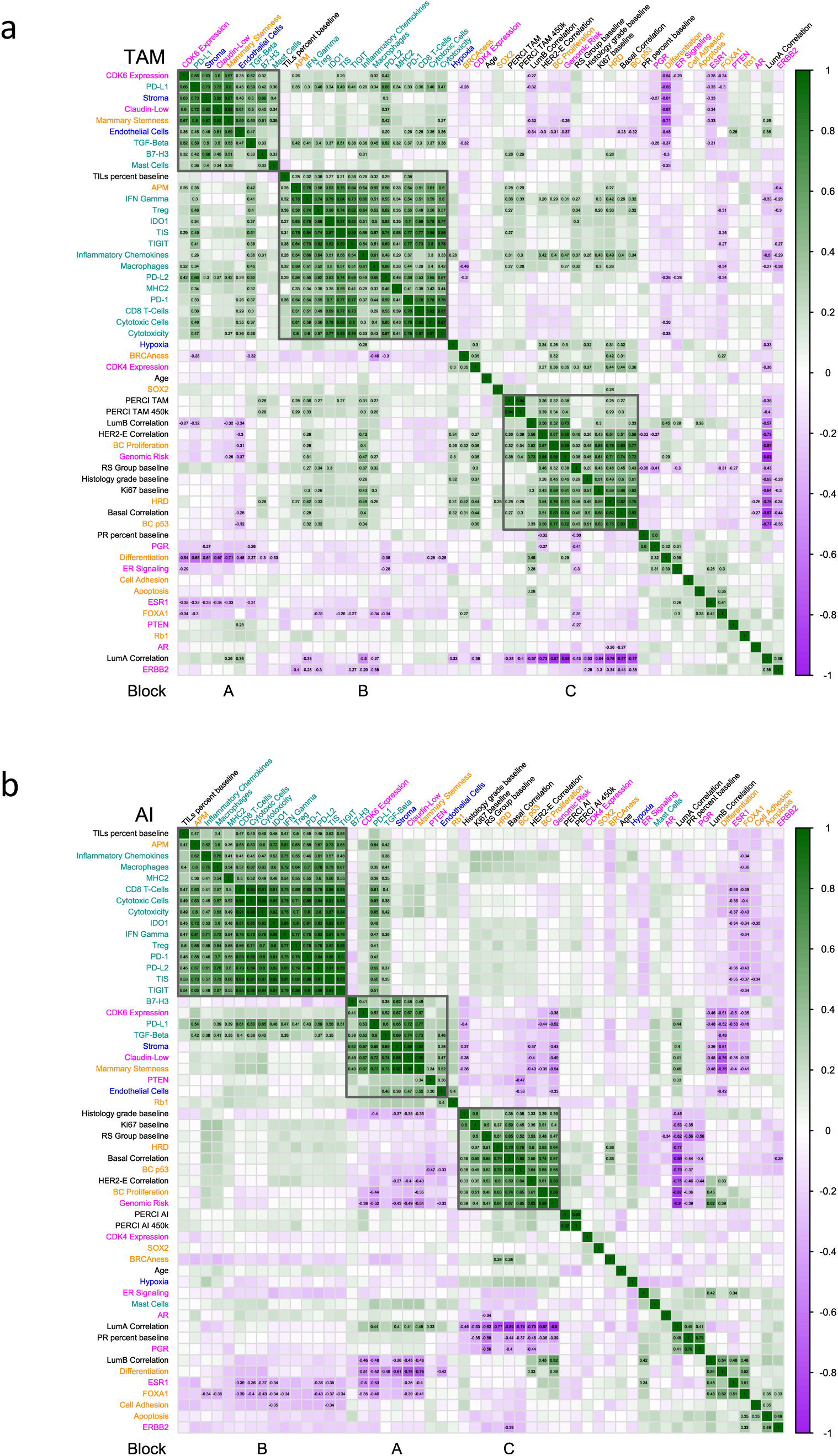
Associations of NanoString signature scores with clinico-pathological parameters and PAM50 correlations in the NanoString cohort. Spearman’s correlation coefficients were calculated between NanoString signature scores (color coded by four NanoString categories: tumor (orange), immune (dark cyan), microenvironment (blue) and breast cancer (magenta)), selected clinical features, PERCI, RS score, and correlation values to the PAM50 subtypes (all black) in the TAM (a) and AI group (b) of the NanoString cohort. Correlations are indicated by a color gradient from purple (−1) to green (1). Results with statistically significant differences between response groups are highlighted by numerical correlation coefficients (two-tailed test of significance which compares the observed value of correlation coefficient to its expected value under the null hypothesis (no correlation between the two variables), fdr-adjusted *p* < 0.01). Further details are described in the Methods section.

Block A (“Reactive tumor microenvironment and stemness”) covered a variety of signatures, including those for the immune-checkpoint protein B7-H3, the cell cycle regulator CDK6, the breast cancer subtype Claudin-Low, Endothelial Cells, Mammary Stemness, Mast Cells, PD-L1, PTEN, Stroma, and TGF-Beta. Block A signatures were negatively correlated with the ESR1, FOXA1 and Differentiation signatures.

Block B (“Immune”) combined both pro- and anti-tumorigenic immune signatures. Pro-tumorigenic activity was indicated by signatures such as APM, IDO1, IFN-Gamma, Inflammatory Chemokines, Macrophages, Programmed cell death protein 1 (PD-1), PD-L2, the immunoreceptor TIGIT, Treg, and the Tumor Inflammation Signature (TIS). Anti-tumorigenic immune signatures include CD8 T-Cells, Cytotoxic Cells, Cytotoxicity, and Major histocompatibility complex class II (MHC-II). The percentage of TILs at baseline showed good correlations with the pro-tumorigenic immune response-related signatures in the TAM group (ρ = 0.27 to 0.38) and with all immune-related signatures in the AI group (ρ = 0.4 to 0.54). This indicates concordance between histopathological measures of immune infiltration and molecular signatures of immune activation.

Block C (“Proliferation and genomic risk”) showed high positive correlations between BC p53, BC Proliferation, Genomic Risk, and HRD signatures and clinical and molecular proliferation markers including RS group, histologic grade, baseline Ki67, and Luminal B, HER2-enriched, and Basal-like PAM50 subtype correlations. The Predictive Endocrine Resistance Index (PERCI) was also strongly correlated with Block C components, primarily in the TAM cohort. PERCI was developed using a penalized logistic regression approach, incorporating data on genomic alterations, patient age, tumour microenvironment composition, and differential DNA methylation. It has been shown to accurately classify patients as either responders or non-responders to endocrine therapy, and to predict progression-free survival.^12^

Block B immune signature correlations were stronger in the AI group than in the TAM group. Conversely, Block C proliferation-related signature correlations were stronger in the TAM group, particularly for the simplified PERCI 450k model, which includes age and selected DNA methylation events, and Luminal B PAM50 correlations (Supplementary Fig. S7).

These results emphasize the molecular differences between the menopausal groups.

### Recurrent genomic alterations differ between menopausal and response groups

To identify recurrent genomic alterations (RGA) that drive endocrine therapy resistance and relate to signature scores, we performed NGS panel sequencing on post-pET tumor samples (Supplementary Table S2).

In the NanoString cohort, *PIK3CA*, *GATA3*, *TP53*, and *MAP3K1* were among the most frequently altered genes (44.5%, 22.3%, 17.7% and 15.5%, respectively), with *MAP3K1* mutations and *GATA3* splicing mutations being more prevalent in the TAM than the AI group (Fig. 2a, Supplementary Table S5). Conversely, *ESR1, RAD51C* and *FGF19* alterations (mainly amplifications) were significantly more often detected in the AI group (*p* < 0.05).

**Fig. 2:**
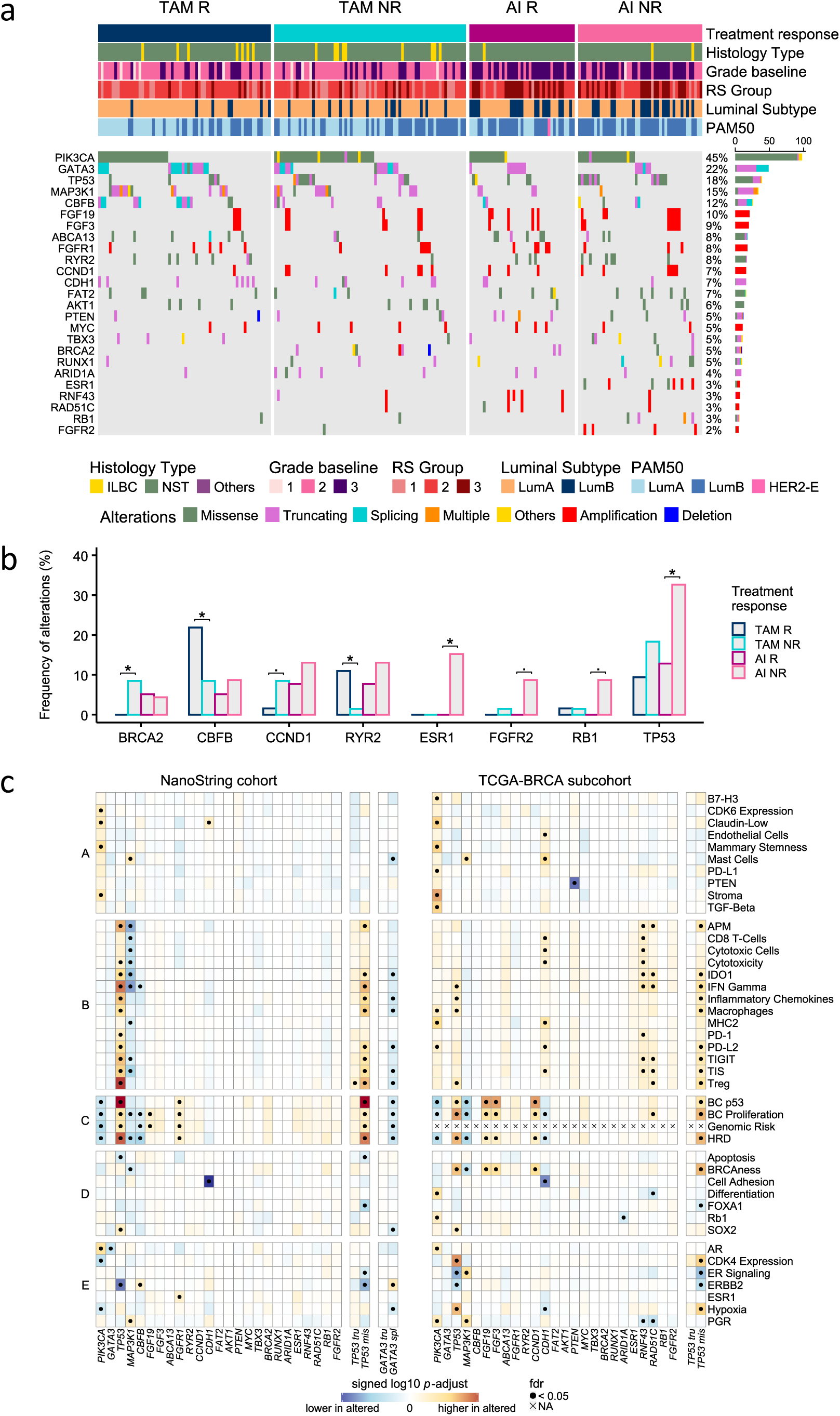
Association of recurrent genomic alterations (RGA) with response and signature scores. (a) The oncoprint summarizes the mutational landscape of top 25 most recurrent genomic alterations, color-coded by the RGA type and separated into TAM R, TAM NR, AI R and AI NR groups. Tumor specimens derived after short-term pET were analyzed for genomic alterations. The barplot at the right quantifies the recurrence of the RGA, and genes are sorted by total alteration burden. Only RGA at minimum 7.5% recurrence in either subgroup are shown. Clinical annotations of cases are indicated on the top. (b) Frequencies of RGA with significant differences between TAM R, TAM NR, AI R and AI NR groups, analyzed using Fisher-exact test with **^⦁^**, *, *p* < 0.1, 0.05. (c) Wilcoxon test for comparison of NanoString signature scores between wild type samples and samples with alterations in the NanoString cohort (n = 220, left) and the TCGA-BRCA subcohort (n = 260, right). For the TCGA-BRCA subcohort, Genomic Risk scores are not available. Color-coded from blue to red according to signed log10 *p*-adjust; ^●^, fdr < 0.05 as indicated.

In the TCGA-BRCA subcohort, *PIK3CA* (42.5%) and *TP53* mutations (17.0%) were confirmed to be frequently mutated (Supplementary Fig. S8, Supplementary Table S6). *CDH1* mutations (19.2%) were detected more often than in the NanoString cohort (7.3%). *GATA3* splicing was less common; however, *GATA3* mutations were still enriched in pre-vs. postmenopausal cases. Conversely, unlike in the NanoString cohort, *MYC* amplifications and *MAP3K1* mutations were more prevalent in post-vs. premenopausal cases (*p* < 0.05).

In the TAM group, *BRCA2* (*p* < 0.05) and *CCND1* (*p* < 0.1) exhibited notably higher alteration frequencies in non-responders than in responders (Fig. 2b). Truncating mutations or deletions of *BRCA2* resulted in significantly lower BRCA2 mRNA expression, while *CCND1* amplifications were associated with significantly higher CCND1 transcript levels (Supplementary Fig. S9). Consistent with our previous findings,^12^ *CBFB* and *RYR2* were more frequently mutated in TAM responders than in non-responders.

The AI non-responder group showed a significant enrichment of *ESR1*, *FGFR2*, *RB1* and *TP53* alterations (Fig. 2b). *ESR1* and *FGFR2* mRNA expression levels were significantly increased (*p* < 0.01) in baseline samples of cases with amplifications of these genes. Truncating *RB1* mutations were associated with decreased RB1 mRNA expression. Similarly, *TP53* gene truncations, which were observed in both treatment groups, were consistently associated with decreased TP53 mRNA levels, indicating the loss of function of the tumor suppressor gene (Supplementary Fig. S9).

### Recurrent genomic alterations impact BC360 signature scores

To functionally associate RGAs with BC360 signatures, we compared the signature scores of wild-type and altered samples for the top 25 RGAs (Fig. 2c, Supplementary Table S2).

In both cohorts, *PIK3CA1* mutations were positively associated with selected Block A (Claudin-Low, Mammary Stemness, Stroma) and AR (Block E) signature scores, while proliferation-related (Block C), CDK4 expression and Hypoxia (both Block E) signature scores were diminished in mutant cases. A robust association was identified between *TP53* mutations, particularly missense mutations, and elevated scores of pro-tumorigenic immune-related (Block B), proliferation-related (Block C), SOX2 (Block D) and hypoxia (Block E) signatures. Similarly, amplifications of chr11q13.3 (*FGF3*, *FGF19*, *CCND1*), chr17q22 (*RAD51C, RNF43*), and chr8p11.23 (*FGFR1*) were associated with increased Block C proliferation signature scores. The amplifications on chr17q22 were inversely related to PRG expression scores.

In contrast to *TP53* mutations, recurrent *MAP3K1* mutations were reproducibly associated with lower immune, proliferation, and BRCAness (Block D) signature scores. Conversely, Mast Cell (Block A) and PGR expression scores (Block E) were higher in mutant cases. The strongest negative association was found between *CDH1* truncating mutations, resulting in significantly reduced *CHD1* mRNA levels (Supplementary Fig. S9), and Cell Adhesion scores. Positive associations of *CDH1* mutations and chr17q22 (*RAD51C, RNF43*) amplifications with pro- and/or anti-tumorigenic immune-related signatures were only detectable in the TCGA-BRCA subcohort.

These findings further support the idea that specific genomic alterations have distinct functional consequences on signature scores, offering mechanistic insights into the molecular heterogeneity associated with primary endocrine resistance.

### Proliferation and pro-tumorigenic immune signatures are elevated in non-responder groups

After relating signature scores to menopause status and RGA, we were interested in whether baseline transcriptional patterns could discriminate treatment-response groups. To that end, we performed a differential signature analysis (Supplementary Table S2).

In the TAM group (Fig. 3a), we identified several pro-tumorigenic immune and proliferation signatures as significantly (fdr < 0.05) upregulated in non-responders (log2FC 0.19-0.54). The finding of elevated BC p53 scores in non-responders was consistent with our previous results, which linked *TP53* alterations and expression to treatment resistance.^21,22^ Conversely, high ERBB2 signature scores were associated with a favorable response to TAM treatment.

**Fig. 3:**
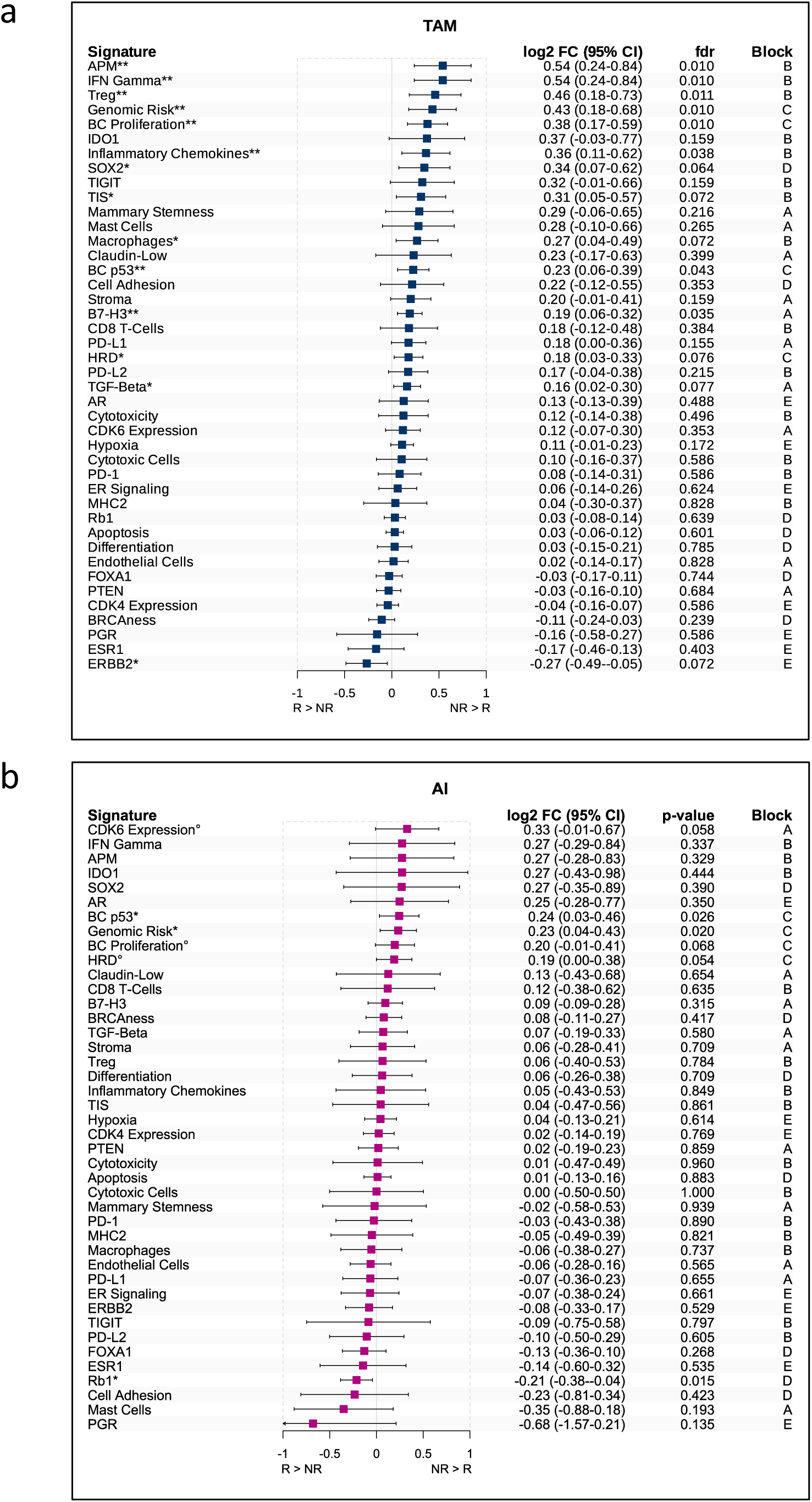
Differential expression of Nanostring signatures between response groups in the NanoString cohort. Summary of differential NanoString signature scores between response groups in the NanoString TAM (a) and AI (b) group by fitting linear mixed models with patient pair_id as a random effect. The following variables are used as covariates: TAM: age group and TILs group at baseline; AI: age group. Signatures are sorted by log2 fold change of mean signature expression between responders and non-responders per treatment. Significant signatures are indicated by °, *, ** with *p*-values < 0.1, 0.05, 0.01 (fdr < 0.05 for TAM). 95% confidence intervals, (fdr-adjusted) *p*-values and block membership are shown.

In the AI group, mainly Block C and CDK6 expression signatures were significantly (*p* < 0.1) higher in non-responders (log2FC 0.2-0.33). In contrast, the Rb1 signature (Block D) was associated with better treatment response (log2FC −0.21) (Fig. 3b).

We further stratified the patients by RS risk groups (Supplementary Fig. S10, Supplementary Table S2). In the low-risk RS1 group enriched in cases from the TAM group, Genomic Risk and BC Proliferation signatures as well as ER Signaling (log2FC 0.37) were significantly (*p* < 0.05) upregulated in non-responders. In the intermediate-risk group RS2.1 of mainly premenopausal women (age ≤ 51), Block A signatures, including Mast cells (log2FC 0.93) and Mammary Stemness signatures (log2FC 0.65), pro-tumorigenic Block B, and BC p53 signatures were significantly higher in the non-responder group, whereas high Differentiation, ESR1 and ERBB2 signature scores indicated better response (log2FC −0.26 - −0.42). In contrast, the intermediate-risk group RS2.2 of older women (age > 51 years) and the high-risk group RS3 exhibited few significant differences between the response groups, suggesting limited discrimination based on signatures in these RS-stratified groups. High SOX2 (log2FC 0.79) and CDK6 (log2FC 0.48) signature scores indicated a worse response in risk group RS2.2, whereas high PD-1 (log2FC −0.58) scores were suggestive of a better response in the RS3 group.

Together, these results demonstrate that distinct transcriptional programs, particularly those related to pro-tumorigenic immune activation and proliferation, exist between treatment response groups. Their relevance may vary by RS and menopausal groups. These findings support the use of gene signatures to understand endocrine response heterogeneity.

### Integration of molecular and genomic features *via* PLS-DA reveals key contributors to patient stratification

To identify the most discriminatory features for patient stratification, we performed a Partial Least Squares Discriminant Analysis (PLS-DA) on the NanoString signature scores, selected RGA and age (Fig. 4). In the TAM group, the combined variables stratified the response groups with reasonable area under the receiver operating curve (ROC-AUC) values (component 1: 73.1%; component 2: 69.2%) (Fig. 4a,b). We performed a feature selection with 50x 5-fold cross validation. The most stable features related to the first two components included proliferation-related signatures (BC proliferation, Genomic Risk, HRD), immune signatures (APM, B7-H3), and PAM50 correlations (HER2-E, LumA). *CBFB* mutations were identified as the most discriminatory RGA (Fig. 4c). The AI group showed a high level of stratification between the response groups, with high AUC values (component 1: 85.9%; component 2, 71.1%) (Fig. 4d,e). The main factor distinguishing between responders and non-responders was age. RGA of *ESR1*, *TP53* and *TBX3* had an important role in stratifying the response groups in component 1, beside HER2-E correlations and signature scores of Rb1, CDK6 Expression (component 1) and B7-H3 and Hypoxia (component 2) (Fig. 4f).

**Fig. 4:**
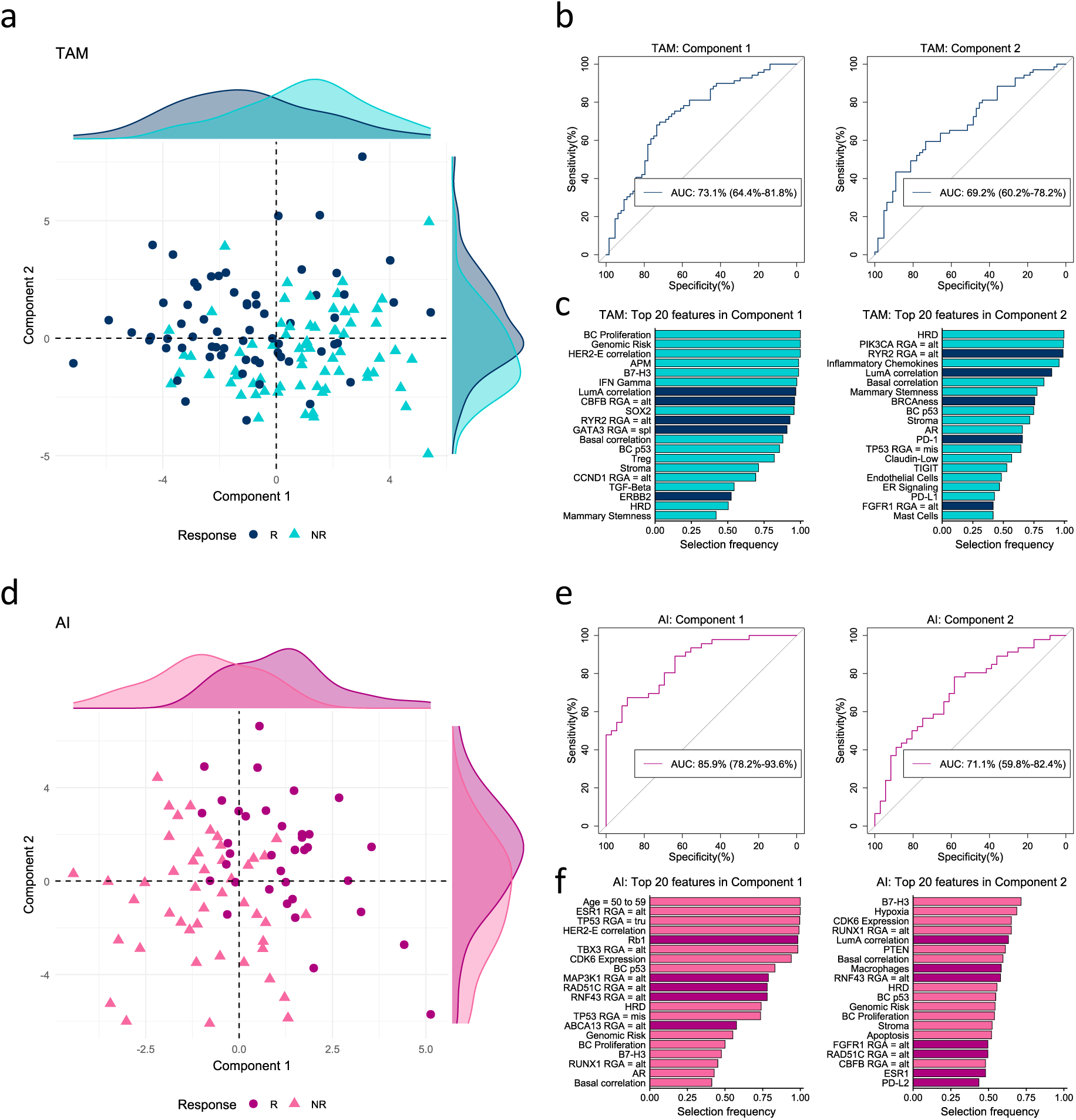
Partial Least Squares Discriminant Analysis (PLS-DA) (a, d) Distribution of samples in component 1 vs. component 2 in the NanoString TAM (a) and AI group (d) based on signature scores, selected RGAs and age in a two-dimensional space using PLS-DA. Responders (R) and non-responders (NR) are indicated by different colors and shapes. Density curves at the outer edges of the plots indicate the number of patients. Components are labeled on the axes. (b, e) Area under the receiver operating characteristic curve (ROC-AUC) of features contributing to component 1 (left) and component 2 (right) to stratify the response groups in the TAM (b) and AI groups (e). (c, f) The top 20 features selected in 50 repetitions of 5-fold cross validation to components 1 and 2 in the TAM (c) and AI (f) groups. The colors in (c) and (f) indicate the group (R or NR) with the higher mean value.

Identifying distinct factors as the most stable discriminators between responders and non-responders in the two menopausal groups suggests differences in underlying resistance mechanisms.

### Baseline NanoString signature scores are associated with long-term survival outcomes

We examined the prognostic significance of baseline transcriptional profiles by linking BC360 signature scores to invasive (IDFS) and distant disease-free survival (DDFS). The median follow-up duration for our cohort was 59.8 months, and follow-up data were available for 191 of the 220 patients included in the study.

Due to the low number of events in the TAM group (Supplementary Fig. S11), we limited these analyses to the AI group. The BC p53 signature score (HR = 2.74 [1.08-6.94]) was the strongest prognosticator for IDFS, whereas *TP53* mutations alone were not prognostically significant (data not shown). Additionally, Genomic Risk (HR = 2.5 [1.07-5.83]), HRD (HR = 2.44 [1.12-5.29]), and Hypoxia (HR = 1.99 [1.12-3.51]) signature scores were associated with worse IDFS (Fig. 5a). Poor survival outcomes for DDFS were linked to higher ERBB2 (HR = 2.46 [1.05-5.75]) and Hypoxia (HR = 2.12 [1.17-3.87]) signature scores. High scores of PD-1 indicated better outcomes (0.44 [0.21-0.94]) (Fig. 5b).

**Fig. 5:**
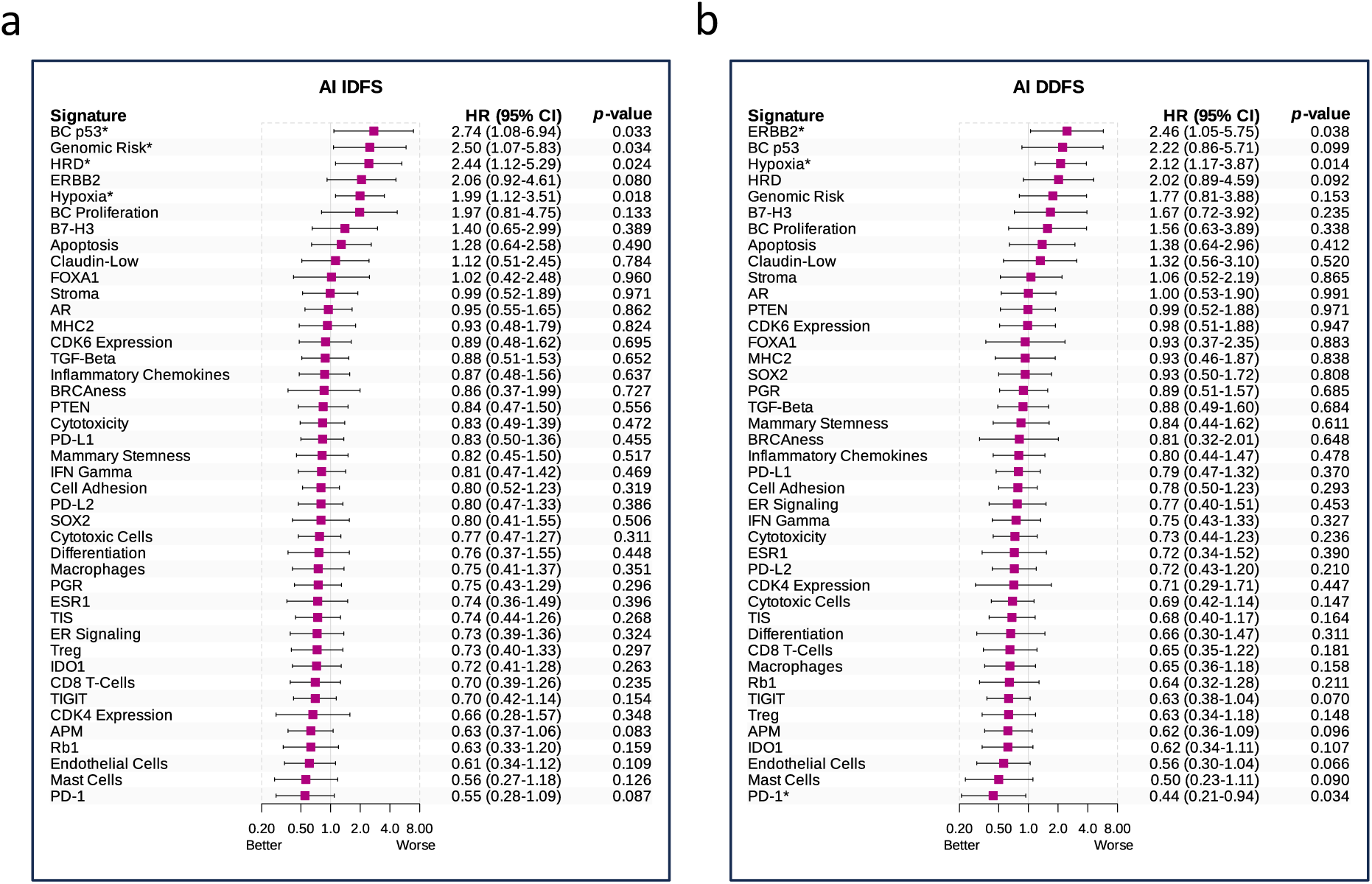
Associations between BC360 signatures and disease-free survival in the AI group. Association of NanoString signatures with survival endpoints (a) invasive disease-free survival (IDFS) and (b) distant disease-free survival (DDFS) in the NanoString AI group were estimated by univariate Cox regression and expressed as hazard ratios (HR) with 95% confidence intervals (95% CI) and *p*-values. Significant signatures are indicated by *, with *p*-value < 0.05. For IDFS, 9 events, and for DDFS, 6 events were recorded (n = 66). Median follow-up duration was 59.8 months.

We used the online tool KM Plotter^20^ to externally validate the clinical relevance of the signatures. Hypoxia, HRD and BC p53 signatures were consistently associated with an increased risk of recurrence and/or distant metastases, and high PGR levels indicated better recurrence-free, metastasis-free and overall survival, confirming our short term response results. Better overall survival was also predicted by higher PD-1 levels (Supplementary Table S7). However, we could not confirm the prognostic relevance of ERBB2 signature scores, which showed reduced rather than increased risk of recurrence or distant metastases. These inconsistent findings may indicate efficient treatment by targeted therapies in the cohorts included in the KM Plotter database.

Together, these findings suggest that certain NanoString signatures at baseline have prognostic value beyond indication of short-term treatment response and may help identify patients at risk for long-term recurrence.

## Discussion

In this study, we employed an integrative approach that combined transcriptomic, genomic, and clinico-pathological features of luminal breast cancer to identify molecular features associated with menopausal status, endocrine response, and long-term survival at baseline. We used the NanoString cohort as a discovery cohort and a subset of the TCGA-BRCA cohort to verify findings related to menopausal status. This was necessary because the NanoString cohort had a partly matched design and was skewed towards high-grade cases in the postmenopausal group. On the other hand, the TCGA-BRCA subcohort had a higher rate of cases classified as ILBC and Luminal A subtype, as well as of lower grade and with equivocal HER2 staining. Our analysis focused on identifying associations that demonstrated consistent trends across both cohorts.

Signature-level profiling of baseline expression showed greater variability in the postmenopausal groups, suggesting increased biological heterogeneity in older patients. A differential signature expression analysis revealed a consistent increase in ESR1 and, to a lesser extend, FOXA1 signature scores and a decrease in signature scores of PTEN and the estrogen-sensor PGR in postmenopausal women, confirming a recent report.^23^ Loss of PGR expression in postmenopausal BC has been associated with endocrine resistance.^24–26^

We also compared mutation frequencies between the menopausal groups. In the NanoString cohort, *GATA3* and *MAP3K1* mutations were more common in premenopausal cases, and mutations in both genes have been associated with better survival in the METABRIC study.^27^ Consistent with the higher prevalence of ILBC cases in the TCGA-BRCA subcohort, *CDH1* mutations were enriched, especially in postmenopausal cases, which also harbored more *MAP3K1* mutations. This finding aligns with the established link between mutant *MAP3K1* and the Luminal A subtype.^28^

Consistent with an earlier study^29^, unsupervised clustering of signature scores resulted in three blocks of highly correlating signatures, which were enriched for reactive stroma and mammary stemness-like features (Block A), immune-related signatures (Block B) and proliferation- and genomic risk-related signatures (Block C). Some of the most frequent RGAs in ER+ breast cancer had distinct associations with signature blocks.^30^ *PIK3CA* mutations were associated with lower proliferation signatures (Block C) and with higher Mammary Stemness scores (Block A), indicating tumors with higher expression of epithelial-to-mesenchymal transition (EMT)-related SNAIL-, ZEB-, and TWIST-family transcription factors. Oncogenic PIK3CA signaling activation has been shown previously to regulate EMT and cancer stemness.^31,32^ We also confirmed an association between *PIK3CA* mutations and elevated AR expression and signaling.^33^ While *PIK3CA* mutations often confer a favorable prognosis, a recent study identified an increased risk of recurrence in high-risk patients with *PIK3CA* mutations (defined by Oncotype DX-, MammaPrint-, and PAM50-ROR-like scores).^34,35^

Both *TP53* missense and truncating mutations were strongly correlated with higher immune- and proliferation-related signature scores (Block B, C), in agreement with our recent work.^21,22^ Similarly, amplifications of *CCND1* on chr11q13.3 suggest a potential resistance mechanism involving deregulated cell cycle progression that overrides the growth-inhibitory effects of endocrine therapy.^36^ In contrast, we found an association between *GATA3* splicing mutations, and lower pro-tumorigenic immune and proliferation signature scores. *GATA3* splice site mutations have been shown to be more prevalent in Luminal A tumors and to be associated with favorable prognosis^37^. Also, *MAP3K1* mutations were associated with lower immune signature scores (Block B), consistent with earlier findings.^38^ These results emphasize that combining genomic and transcriptomic data provides a more comprehensive understanding of the potential mechanisms that drive endocrine resistance than using clinical markers or single assays alone.

Differential signature expression analysis between treatment-response groups (Fig. 3a) and PLS-DA (Fig. 4a-c) reinforced the significant role of proliferation and immune-related programs within the TAM group, where non-responders exhibited higher expression of Block B and C signatures. This aligns with studies showing that high proliferation^39^ and specific immune profiles^40^ can be associated with tamoxifen resistance and poor prognosis.^41^ Additional important features discriminating R and NR groups were PAM50 LumA and HER2 correlation, and *CBFB* mutations. *CBFB* loss of function mutations lead to metabolic reprogramming and have recently been shown to synergize with *PIK3CA* gain of function mutation to promote breast tumor progression.^42^ Interestingly, the AI group displayed fewer significant signature score differences (Fig. 3b), which may stem from both biological factors, such as more heterogeneous resistance mechanisms within this group^43^, and statistical limitations due to sample size or the magnitude of gene expression changes. The observed broader transcriptomic variability in the AI cohort could also mask consistent resistance signatures changes. According to PLS-DA, the combination of young age, driver mutations in *ESR1, TP53 and TBX3*, HER2 correlation, and CDK6 and Rb1 signature scores provided the most effective discrimination of responders from non-responders within the AI group. *TBX3* is a transcriptional regulator of the ER and was found to be enriched post-pET in endocrine-resistant advanced ER+ BC.^44^

Analysis of baseline transcriptional profiles in relation to long-term outcomes (IDFS and DDFS) in the AI group revealed distinct prognostic associations (Fig. 5). Elevated baseline scores for BC p53, Genomic Risk, HRD, and Hypoxia signatures were associated with worse IDFS. For DDFS, higher ERBB2 and Hypoxia signature scores predicted poor survival. Our short-term resistance analysis in the AI cohort indicated higher expression of BC p53 and Genomic Risk in non-responders, partially mirroring the long-term prognostic associations of these signatures with poorer IDFS. Additionally, the higher frequency of recurrent genomic alterations in *ESR1*, *FGFR2*, *RB1*, and *TP53* observed in AI non-responders may contribute modestly to early resistance but appear more strongly associated with mechanisms underlying subsequent disease recurrence.

Our study provides an explorative analysis of transcriptomic, genomic, and clinico-pathological features associated with the response to short-term endocrine therapy and the long-term prognosis of luminal breast cancer. Although decision curve analysis (DCA) would be a valid strategy for evaluating the clinical utility of our results as predictive or prognostic models,^45^ due to the limitations in our study design outlined below, we refrained from developing a clinical decision model. Nevertheless, the clinical relevance of our findings can be discussed in the context of clinical trial results.

Block A signatures scores including the CDK6 signature were negatively correlated with the Differentiation signature score, and CDK6 scores were elevated in non-responders of the AI group. Elevated CDK6 can likely bypass the growth-inhibitory effects of endocrine therapy.^46^ The MONALEESA-2 trial reported progression-free and overall survival benefits of the CDK4/6 inhibitor ribociclib in combination with AI in postmenopausal HR+ HER2-advanced breast cancer independent of RGA status.^47^ The success of CDK4/6 inhibitors, which are suggested to act both on tumor cells and on the tumor immune environment, highlights the complex interplay between intrinsic tumor properties (differentiation state) and extrinsic factors (microenvironment) in dictating therapeutic response.^48^

Mutant TP53, particularly missense mutations, is associated with elevated pro-tumorigenic Block B immune signature scores. In the TAM cohort, various immune-related signature scores from Block B were higher in non-responders. Immune checkpoint inhibitors have primarily been tested as a combination therapy for triple-negative and HER2-overexpressing BC. Results from clinical trials of early-stage luminal BC indicate that combination therapy could be beneficial for high-risk luminal B-like tumors. While the findings are encouraging, there is a clear need to develop biomarkers that can effectively predict treatment response, resistance, and immune-related toxicities.^49–53^ Since TP53 mutations lead to aberrant nuclear accumulation of the mutant p53 protein,^54^ p53 IHC may potentially be used as a surrogate marker of endocrine resistance in a clinical setting and identify cases for whom the use of adjuvant chemo- or chemo-immunotherapy would be justified.^21,50^

We observed a strong negative correlation between the Luminal A subtype and proliferation-related Block C signatures, which were significantly higher at baseline in non-responders in both treatment groups as well as in all RS groups when analyzed separately. These findings support the established understanding that tumors with higher Luminal A characteristics and low baseline proliferation rates are associated with greater endocrine responsiveness.^55^ BC Proliferation and Genomic Risk signature scores indicate aggressive disease and correlate highly with Ki67 staining and RS grouping. The use of RS to inform treatment decisions has undergone testing in clinical trials, indicating a benefit of chemo-endocrine therapy for younger patients in the intermediate-risk group RS2.^56,57^

Finally, PGR scores (Block E) were significantly higher in premenopausal cases than in postmenopausal cases, and had a trend of being higher expressed in responders than in non-responders. High PGR signature scores in the TAM cohort were indicative of good prognosis. Our external validation supports high PGR expression as an indication of good prognosis,^58–61^ suggesting PGR expression as a marker for risk stratification.

Our study has limitations that should be considered when interpreting the findings. The cohort size, although prospectively defined and clinically annotated, is relatively modest for survival analysis in the TAM cohort with relatively young patients. The statistical approach is explorative and hypothesis generating. Our dataset is derived from a discovery cohort with a matched-pair design. However, there were sample dropouts due to limited RNA availability, resulting in partially matched pairs. Therefore, the study is neither designed nor powered to develop or validate a baseline predictive model of endocrine resistance suitable for guiding treatment decisions. Besides, the NanoString BC360 panel, while enriched for breast cancer relevant biology, covers a limited number of genes and may not capture the full complexity of endocrine resistance. Nevertheless, the ADAPT trial is a worldwide unique phase III trial allowing analysis of factors impacting on endocrine sensitivity in a cohort of pre- and postmenopausal patients receiving either tamoxifen or AI in HR+ HER2-early breast cancer^6^.

## Conclusions

By leveraging short-term endocrine response data from a clinically annotated unique trial cohort, we demonstrate that distinct baseline molecular profiles, particularly involving immune activation, proliferation, tumor differentiation, and specific recurrent genomic alterations, are predictive of endocrine resistance.

Key findings of our hypothesis generating approach include the association of high proliferation and immune-related gene signature scores with TAM resistance, and the enrichment of *TP53* mutations and amplifications of chr11q13.3 (*FGF3*, *FGF19*, *CCND1*) among non-responders. These alterations were not only more frequent in resistant tumors but also reflected functional consequences at the transcriptomic level. Our explorative results underscore the importance of combining transcriptomic and genomic information for improved patient stratification and suggest that early transcriptional and mutational signatures may inform both therapy selection and long-term outcome predictions. These insights could be validated in future studies to support efforts to develop more personalized endocrine therapy strategies and lay the foundation for integrating multi-omic biomarkers into clinical decision-making for early-stage ER-positive breast cancer.

## List of abbreviations

ADAPT: adjuvant dynamic marker-adjusted personalized therapy
AI: aromatase inhibitors
APM: Antigen processing machinery
AUC-ROC: Area Under the Receiver Operating Characteristic Curve
BC: breast cancer
BC360: NanoString Breast Cancer 360™ gene expression panel
CI: Confidence Interval
DDFS: distant disease-free survival
ER: estrogen receptor
FDR: false discovery rate
FFPE: formalin-fixed paraffin-embedded
HER2: human epidermal growth factor receptor 2 (official name: ERBB2)
HR: hazard ratio
HRD: homologous recombination deficiency
IDFS: invasive disease-free survival
IHC: immunohistochemistry
ILBC: Infiltrating lobular breast cancer
IQR: interquartile range
Ki67: proliferation marker Ki67
MHC: major histocompatibility complex
NGS: next-generation sequencing
NR: non-responder
PD-1: programmed cell death protein 1
PERCI: Predictive Endocrine ResistanCe Index
pET: preoperative endocrine therapy
PGR: progesterone receptor
PLS-DA: Partial Least Squares Discriminant Analysis
pN: pathologic lymph node status
PR: progesterone receptor
pT: pathologic tumor stage
R: responder
RGA: recurrent genomic alteration
ROR: Risk of Recurrence
RS: Recurrence Score
SERMs: selective estrogen receptor modulators
TAM: tamoxifen
TILs: tumor-infiltrating lymphocytes
TIS: Tumor Inflammation Signature
TP53: tumor suppressor protein p53
WSG: West German Study Group

## Ethics approval and consent to participate

The study design is following the guidelines of the local ethics committee (Ethics committee of the Medical School Hannover, ID 2716-2015). Written consent was obtained by the participants.

## Consent for publication

Not applicable.

## Availability of data and materials

The NanoString RNA expression dataset supporting the conclusions of this article is available in the GEO repository under accession number GSE277607. The NGS panel sequencing data will be available in the EGA repository with accession number EGAD50000001595. Access to survival data from the WSG-ADAPT trial is restricted. TCGA-BRCA mutation and copy-number data (PanCancer Atlas, 2018) were obtained from cBioPortal (accessed 2024-02). Normalized PanCanAtlas RNA-seq data were downloaded from <u>gdc.cancer.gov</u> (last accessed 2025-09-17).

## Authors’ contributions

H.K., N.H. and M.C. designed the study. O.G., M.G., S.K., U.N. and N.H. collected patient samples and data. R.K. and M.C. selected the patients and performed the matching. M.C., H.C., L.D.K. and M.R. performed the immunohistochemical stainings and pathology assessment. G.Z., C.G., S.B., H.N., U.M., L.M., and C.zE. performed the experiments, analyzed, and interpreted the data. U.M. provided supervision. G.Z. and C.G. wrote the manuscript. H.K., C.P., N.H. and U.M. reviewed and edited the manuscript. All authors read and approved the final manuscript.

## Data Availability

The NanoString RNA expression dataset supporting the conclusions of this article is available in the GEO repository under accession number GSE277607. The NGS panel sequencing data will be available in the EGA repository with accession number EGAD50000001595. Access to survival data from the WSG-ADAPT trial is restricted. TCGA-BRCA mutation and copy-number data (PanCancer Atlas, 2018) were obtained from cBioPortal (accessed 2024-02). Normalized PanCanAtlas RNA-seq data were downloaded from gdc.cancer.gov (last accessed 2025-09-17).

https://www.ncbi.nlm.nih.gov/geo/query/acc.cgi?acc=GSE277607

https://ega-archive.org/datasets/EGAD50000001595

## Data Availability

https://www.ncbi.nlm.nih.gov/geo/query/acc.cgi?acc=GSE277607

https://ega-archive.org/datasets/EGAD50000001595

## Acknowledgements

We thank the patients and families who contributed to this study.

The results reported here are in part based upon data generated by The Cancer Genome Atlas managed by the NCI and NHGRI. Information about TCGA can be found at http://cancergenome.nih.gov.

## Declaration of Generative AI and AI-assisted technologies in the writing process

Statement: During the preparation of this work the author(s) used ChatGPT (OpenAI) in order to improve readability and language flow in the manuscript. After using this tool/service, the author(s) reviewed and edited the content as needed and take(s) full responsibility for the content of the publication.

