## Supplementary figures for "Integrative analysis of pre-treatment RNA expression signatures and recurrent genomic alterations: Link to menopausal status, short-term endocrine therapy response and disease-free survival in luminal breast cancer"

**Table of contents:**

**Supplementary Fig. S1:** Descriptive statistics of the NanoString cohort.

**Supplementary Fig. S2:** Overview of signature scores in the NanoString cohort.

**Supplementary Fig. S3:** Overview of signature scores in the TCGA-BRCA subcohort.

**Supplementary Fig. S4:** Variance of signature scores in the NanoString and TCGA-BRCA subcohort.

**Supplementary Fig. S5:** Covariate-adjusted differential expression of Nanostring signatures between pre- (TAM) and postmenopausal cases (AI).

**Supplementary Fig. S6:** Correlations of BC360 signature scores in the TCGA-BRCA subcohort

**Supplementary Fig. S7:** Differences in signature score correlations in the NanoString TAM and AI groups.

**Supplementary Fig. S8:** Oncoprint of RGAs in the TCGA-BRCA subcohort.

**Supplementary Fig. S9:** RGAs resulting in altered gene expression in the NanoString cohort.

**Supplementary Fig. S10:** Differential expression of Nanostring signatures between R and NR per RS risk groups.

**Supplementary Fig. S11:** Associations between signatures and disease-free survival in the TAM group.

**Supplementary Fig. S1: Descriptive statistics of the NanoString cohort.**

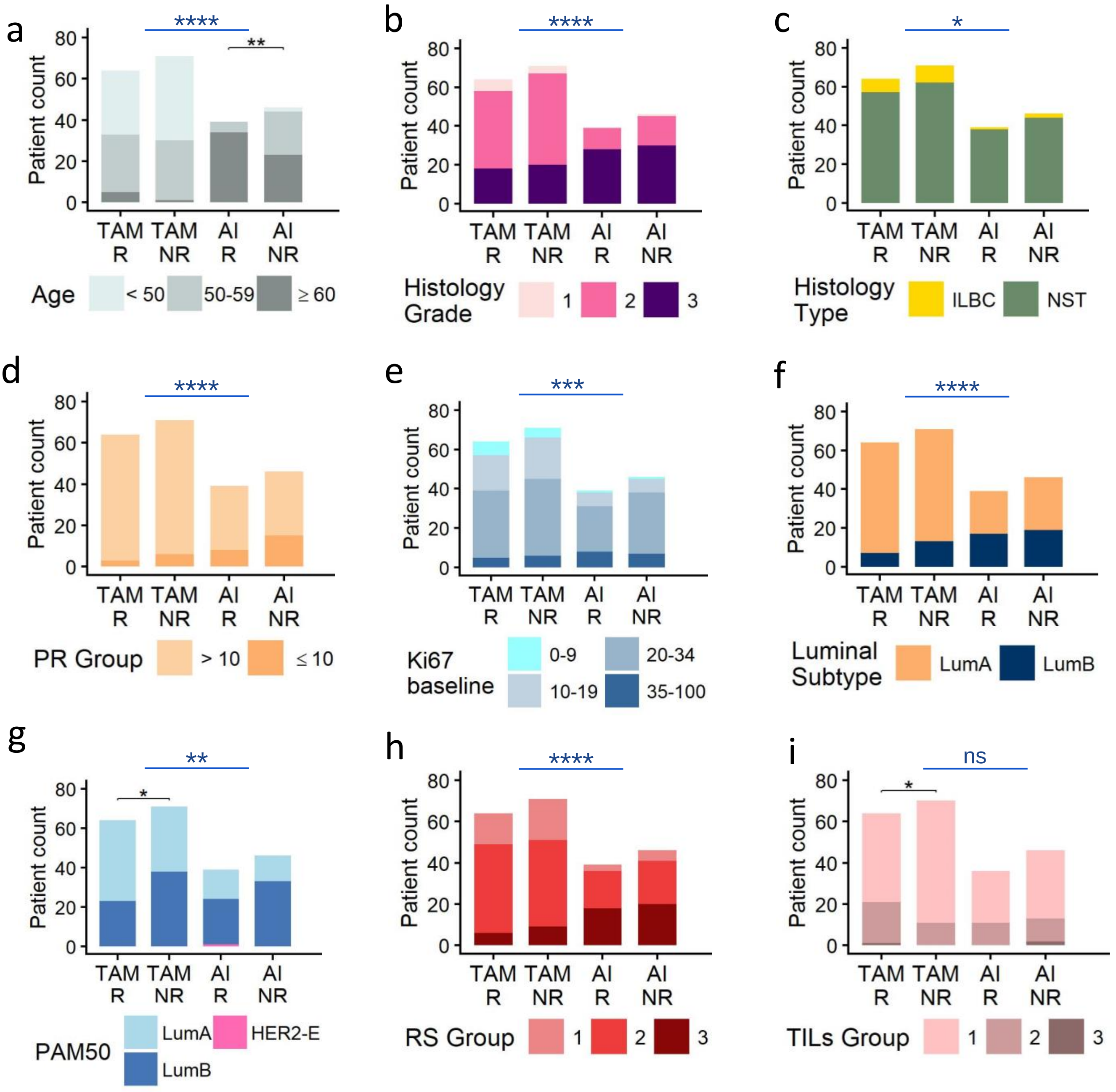


**Supplementary Fig. S1: Descriptive statistics of the NanoString cohort.** Distribution of patients (n=220), analyzed at baseline prior to treatment with TAM in premenopausal (n = 135) or AI in postmenopausal cases (n = 85). Patients are stratified by later response to short-term pET into responder (R) and non-responder (NR) according to: (a) age; (b) histology grade; (c) histology type; (d) progesterone receptor (PR) status; (e) percentage of Ki67-positive staining in IHC at baseline; (f) luminal subtype; (g) intrinsic subtypes of breast cancer using PAM50 gene expression signatures; (h) recurrence score (RS); (i) tumor-infiltrating lymphocytes in pathologic tissue sections (TILs, patients without data are excluded). Comparisons between R and NR cases were performed using a Cumulative Link Mixed Model for ordinal variables and a Generalized Linear Mixed-Effects Model for binary variables with patient pair ID as a random effect where applicable. For sparse data with complete or near separation, Fisher's exact test was applied. Asterisks *, ** indicate p-values < 0.05, 0.01. (a-h) Lines and asterisks *,**,***,**** in blue above the graphs indicate *p*-values < 0.05, 0.01, 0.001, 0.0001 for comparisons between pre- and postmenopausal cases, analyzed using the chi-squared test for trends for Ki67, and Fisher's exact test for all other features. ns: not significant

**Supplementary Fig. S2: Overview of signature scores in the NanoString cohort.**

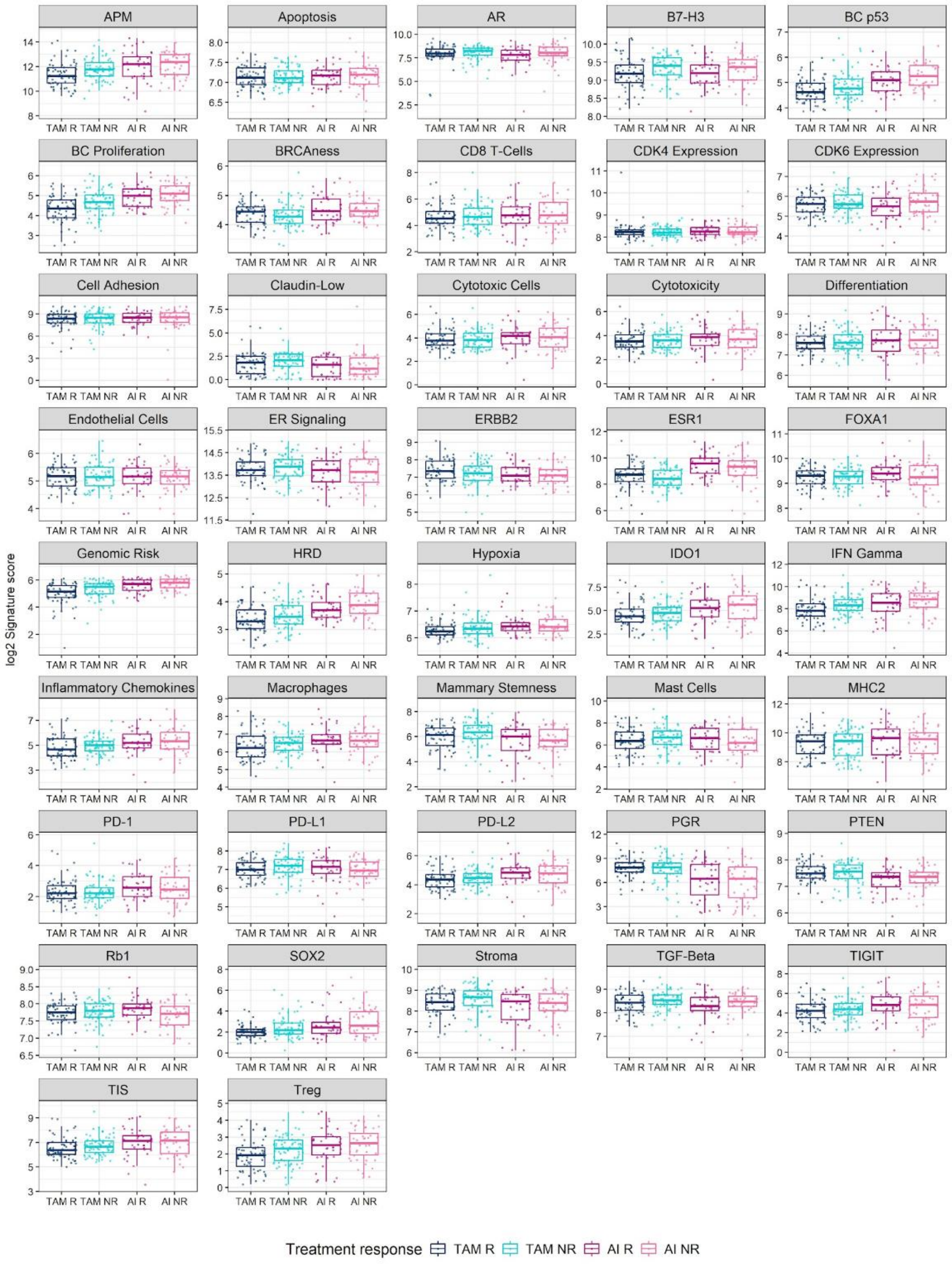

**Supplementary Fig. S2: Overview of signature scores in the NanoString cohort.** NanoString signature scores are stratified by treatment and response groups. Boxplots display the median (line), interquartile range (box), and whiskers extending to 1.5 times the interquartile range (IQR).

**Supplementary Fig. S3: Overview of signature scores in the TCGA-BRCA subcohort.**

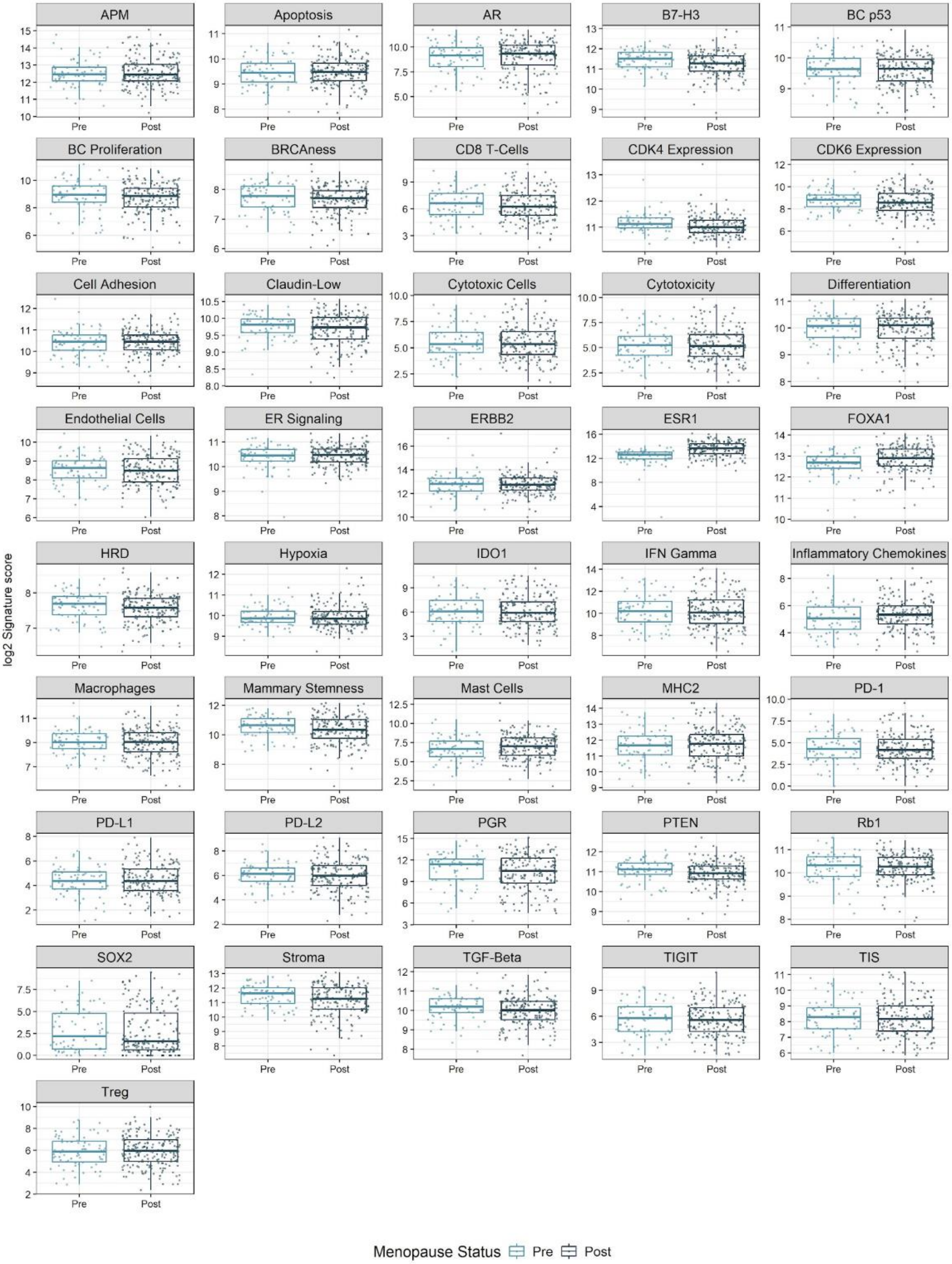

**Supplementary Fig. S3: Overview of signature scores in the TCGA-BRCA subcohort.** Signature scores in the TCGA-BRCA subcohort, stratified by menopausal status. Signature scores were computed as means of log2 mRNA expression values of all genes per signature (with exceptions, see methods). Boxplots display the median (line), interquartile range (box), and whiskers extending to 1.5 times the interquartile range (IQR).

**Supplementary Fig. S4: Variance of signature scores in the NanoString and TCGA-BRCA subcohort**

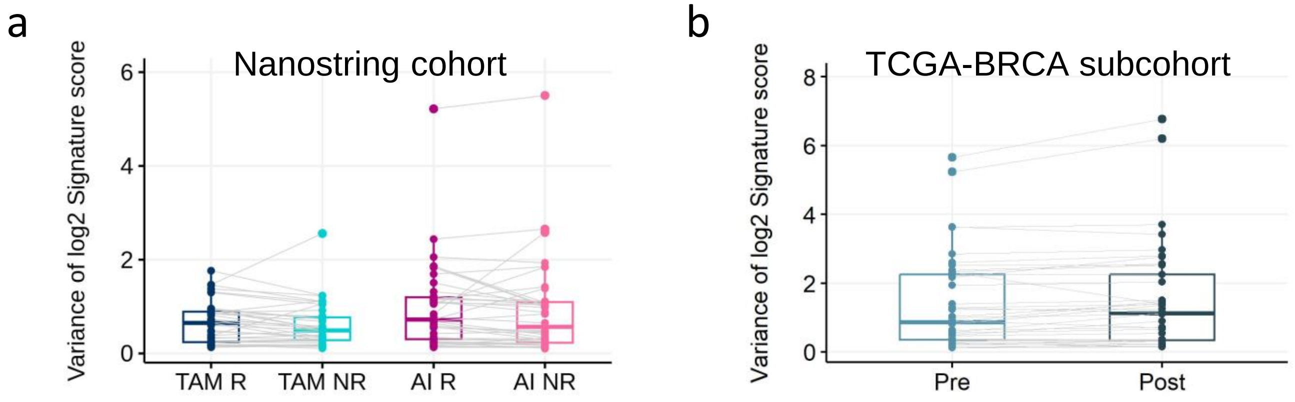


**Supplementary Fig. S4: Variance of signature scores in the NanoString and TCGA-BRCA subcohort**

(a,b) NanoString BC360 signature scores were used to calculate variance (a) within each treatment and response group of the NanoString cohort (a), and within menopausal groups in the TCGA-BRCA subcohort (b). Identical signatures across groups are connected by grey lines. Boxplots display the median (line), interquartile range (box), and whiskers extending to 1.5 times the interquartile range (IQR).

**Supplementary Fig. S5: Covariate-adjusted differential expression of NanoString signatures between pre- (TAM) and postmenopausal cases (AI).**

a

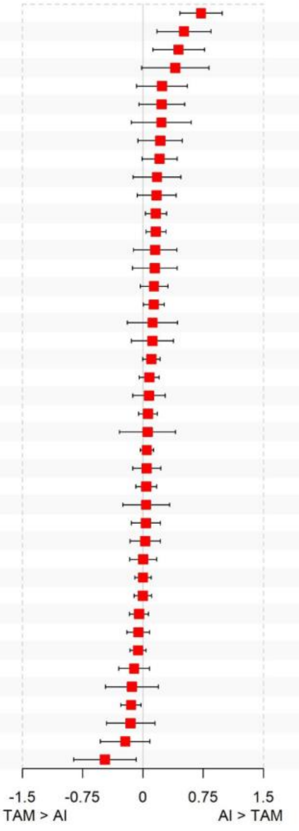


| Signature | log2 FC (95% CI) | *p*-value | fdr |
|---|---|---|---|
| ESR1** | 0.72 (0.46-0.99) | 0.000 | 0.000 |
| SOX2* | 0.51 (0.17-0.85) | 0.003 | 0.066 |
| APM* | 0.44 (0.12-0.76) | 0.007 | 0.097 |
| IDO1 | 0.40 (-0.01-0.82) | 0.058 | 0.245 |
| IFN Gamma | 0.24 (-0.08-0.55) | 0.142 | 0.398 |
| TIS | 0.23 (-0.05-0.52) | 0.107 | 0.368 |
| TIGIT | 0.23 (-0.14-0.60) | 0.222 | 0.469 |
| Treg | 0.22 (-0.06-0.49) | 0.126 | 0.377 |
| PD-L2 | 0.21 (-0.01-0.43) | 0.064 | 0.245 |
| CD8 T-Cells | 0.18 (-0.12-0.47) | 0.243 | 0.469 |
| PD-1 | 0.17 (-0.07-0.41) | 0.168 | 0.416 |
| Genomic Risk* | 0.16 (0.03-0.30) | 0.017 | 0.120 |
| BC Proliferation* | 0.16 (0.04-0.29) | 0.010 | 0.110 |
| Cytotoxicity | 0.15 (-0.12-0.42) | 0.268 | 0.469 |
| Cytotoxic Cells | 0.15 (-0.13-0.43) | 0.296 | 0.481 |
| Differentiation | 0.14 (-0.03-0.31) | 0.114 | 0.368 |
| FOXA1* | 0.14 (0.00-0.26) | 0.042 | 0.223 |
| MHC2 | 0.12 (-0.19-0.43) | 0.450 | 0.586 |
| Inflammatory Chemokines | 0.12 (-0.14-0.38) | 0.373 | 0.559 |
| HRD | 0.10 (0.00-0.22) | 0.059 | 0.245 |
| BRCAness | 0.08 (-0.04-0.20) | 0.207 | 0.469 |
| Macrophages | 0.08 (-0.13-0.28) | 0.461 | 0.586 |
| BC p53 | 0.06 (-0.06-0.18) | 0.298 | 0.481 |
| Mast Cells | 0.06 (-0.29-0.41) | 0.740 | 0.840 |
| Apoptosis | 0.05 (-0.03-0.13) | 0.252 | 0.469 |
| CDK6 Expression | 0.05 (-0.13-0.22) | 0.598 | 0.717 |
| Endothelial Cells | 0.04 (-0.09-0.17) | 0.524 | 0.648 |
| AR | 0.04 (-0.25-0.33) | 0.784 | 0.845 |
| ER Signaling | 0.04 (-0.14-0.22) | 0.687 | 0.801 |
| ERBB2 | 0.03 (-0.16-0.22) | 0.768 | 0.845 |
| PD-L1 | 0.00 (-0.16-0.17) | 0.960 | 0.990 |
| Rb1 | 0.00 (-0.10-0.10) | 0.966 | 0.990 |
| Hypoxia | 0.00 (-0.11-0.11) | 0.994 | 0.994 |
| B7-H3 | -0.05 (-0.17-0.07) | 0.426 | 0.577 |
| TGF-Beta | -0.06 (-0.20-0.08) | 0.425 | 0.577 |
| CDK4 Expression | -0.06 (-0.16-0.04) | 0.230 | 0.469 |
| Stroma | -0.11 (-0.30-0.08) | 0.266 | 0.469 |
| Claudin-Low | -0.14 (-0.46-0.19) | 0.417 | 0.577 |
| PTEN* | -0.15 (-0.27--0.02) | 0.021 | 0.124 |
| Cell Adhesion | -0.15 (-0.45-0.15) | 0.317 | 0.493 |
| Mammary Stemness | -0.22 (-0.53-0.09) | 0.161 | 0.416 |
| PGR* | -0.47 (-0.86--0.09) | 0.017 | 0.120 |

b

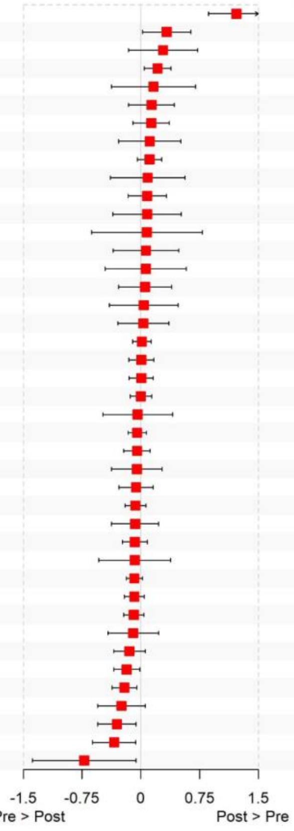


| Signature | log2 FC (95% CI) | *p*-value | fdr |
|---|---|---|---|
| ESR1** | 1.22 (0.86-1.57) | 0.000 | 0.000 |
| Inflammatory Chemokines* | 0.33 (0.02-0.64) | 0.037 | 0.197 |
| Mast Cells | 0.28 (-0.16-0.73) | 0.209 | 0.585 |
| FOXA1* | 0.21 (0.04-0.38) | 0.015 | 0.131 |
| IDO1 | 0.16 (-0.38-0.70) | 0.557 | 0.931 |
| MHC2 | 0.14 (-0.16-0.43) | 0.360 | 0.797 |
| APM | 0.13 (-0.10-0.36) | 0.262 | 0.687 |
| Treg | 0.11 (-0.28-0.51) | 0.574 | 0.931 |
| Apoptosis | 0.11 (-0.05-0.27) | 0.165 | 0.552 |
| PD-1 | 0.09 (-0.39-0.56) | 0.722 | 0.934 |
| ERBB2 | 0.08 (-0.16-0.32) | 0.516 | 0.931 |
| Cytotoxicity | 0.08 (-0.36-0.52) | 0.723 | 0.934 |
| SOX2 | 0.08 (-0.63-0.78) | 0.833 | 0.934 |
| IFN Gamma | 0.06 (-0.35-0.48) | 0.759 | 0.934 |
| TIGIT | 0.06 (-0.45-0.58) | 0.809 | 0.934 |
| PD-L1 | 0.05 (-0.29-0.39) | 0.756 | 0.934 |
| Cytotoxic Cells | 0.04 (-0.40-0.48) | 0.867 | 0.934 |
| TIS | 0.03 (-0.29-0.36) | 0.841 | 0.934 |
| ER Signaling | 0.01 (-0.10-0.13) | 0.831 | 0.934 |
| Cell Adhesion | 0.01 (-0.16-0.17) | 0.944 | 0.969 |
| Hypoxia | 0.00 (-0.15-0.16) | 0.946 | 0.969 |
| BC p53 | 0.00 (-0.14-0.14) | 0.998 | 0.998 |
| AR | -0.04 (-0.49-0.40) | 0.857 | 0.934 |
| HRD | -0.05 (-0.16-0.07) | 0.412 | 0.864 |
| Differentiation | -0.05 (-0.22-0.12) | 0.563 | 0.931 |
| Macrophages | -0.05 (-0.37-0.27) | 0.752 | 0.934 |
| Endothelial Cells | -0.06 (-0.28-0.16) | 0.576 | 0.931 |
| BRCAness | -0.07 (-0.20-0.06) | 0.304 | 0.751 |
| BC Proliferation | -0.07 (-0.37-0.23) | 0.630 | 0.934 |
| Rb1 | -0.08 (-0.24-0.08) | 0.346 | 0.797 |
| CD8 T-Cells | -0.08 (-0.54-0.38) | 0.737 | 0.934 |
| CDK4 Expression | -0.08 (-0.19-0.02) | 0.118 | 0.494 |
| Claudin-Low | -0.08 (-0.21-0.04) | 0.188 | 0.564 |
| Genomic Risk | -0.09 (-0.22-0.04) | 0.171 | 0.552 |
| PD-L2 | -0.10 (-0.42-0.22) | 0.551 | 0.931 |
| TGF-Beta | -0.15 (-0.35-0.06) | 0.155 | 0.552 |
| PTEN* | -0.18 (-0.35--0.01) | 0.033 | 0.197 |
| B7-H3* | -0.21 (-0.37--0.05) | 0.009 | 0.131 |
| CDK6 Expression | -0.25 (-0.55-0.06) | 0.109 | 0.494 |
| Mammary Stemness* | -0.31 (-0.55--0.06) | 0.015 | 0.131 |
| Stroma* | -0.34 (-0.62--0.06) | 0.016 | 0.131 |
| PGR* | -0.72 (-1.38--0.06) | 0.032 | 0.197 |

**Supplementary Fig. S5: Covariate-adjusted differential expression of NanoString signatures between pre- (TAM) and postmenopausal cases (AI).** (a, b) Summary of differential NanoString signature scores between TAM and AI groups in the NanoString cohort (a) and pre- and postmenopausal groups in the TCGA-BRCA subcohort (b), by fitting linear mixed models. We adjusted for clinical characteristics with significant differences between menopausal groups (except for age): (a): Histological grade, histological subtype, PR percent baseline (grouped), Ki67 baseline (grouped), luminal subtype, PAM50, E-cadherin baseline and RS groups; (b): histological stage and race. Signatures are sorted by log2 fold change of mean signature expression. 95% confidence intervals of log2 fold change, *p*-values and fdr-adjusted *p*-values are shown. Significant signatures are indicated by *, *p*-values < 0.05; **, fdr-adjusted *p*-values < 0.05.

### Supplementary Fig. S6: Correlations of BC360 signature scores in the TCGA-BRCA subcohort

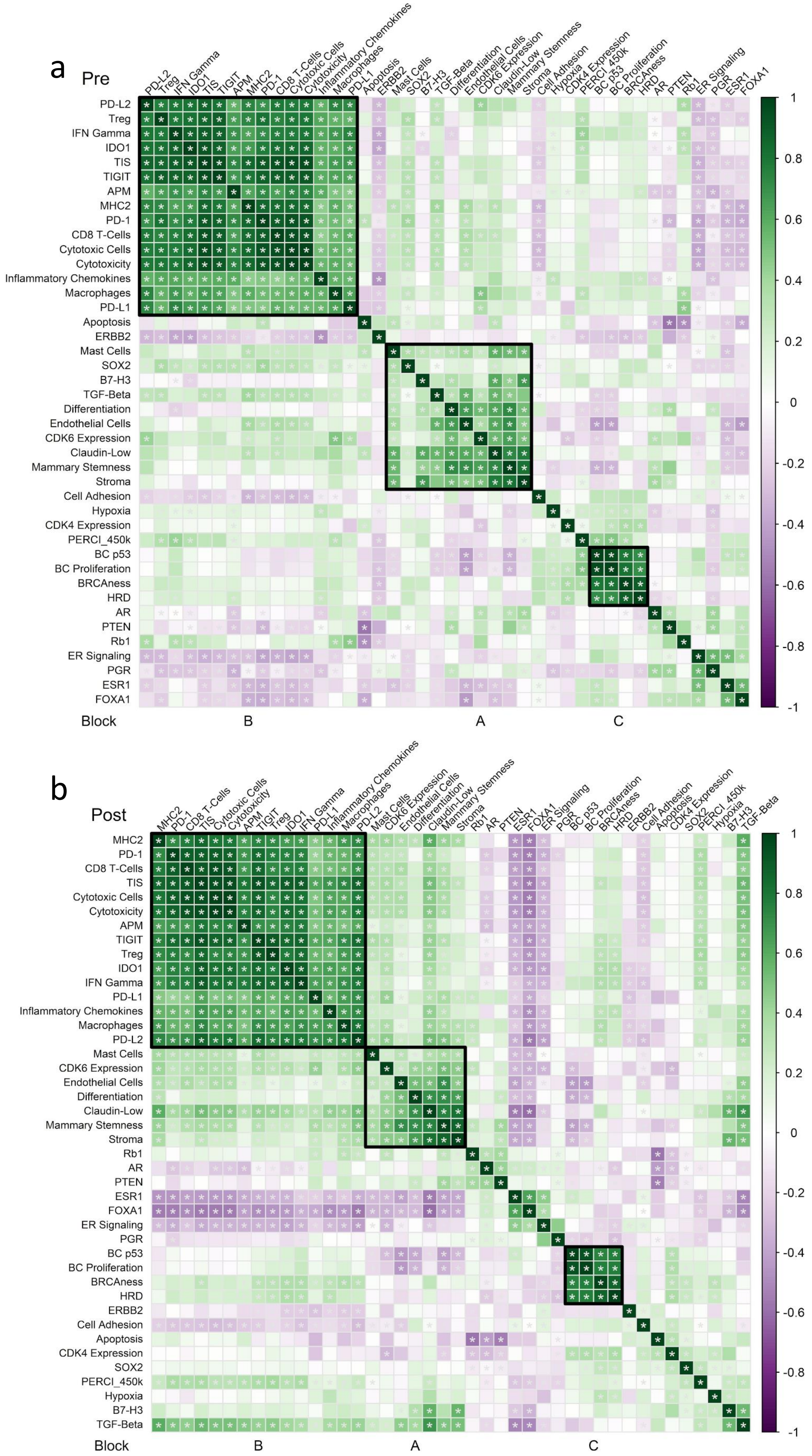

**Supplementary Fig. S6: Correlations of BC360 signature scores in the TCGA-BRCA subcohort**

Spearman's correlation coefficients were calculated between NanoString signature scores and PERCI-450k in premenopausal (a) and postmenopausal cases (b) of the TCGA-BRCA subcohort. Correlations are indicated by a color gradient from purple (-1) to green (1). Results with statistically significant differences between response groups are indicated by an asterisk (two-tailed test of significance which compares the observed value of correlation coefficient to its expected value under the null hypothesis (no correlation between the two variables), fdr-adjusted $p$-value < 0.01).

**Supplementary Fig. S7: Differences in signature score correlations in the NanoString TAM and AI groups**

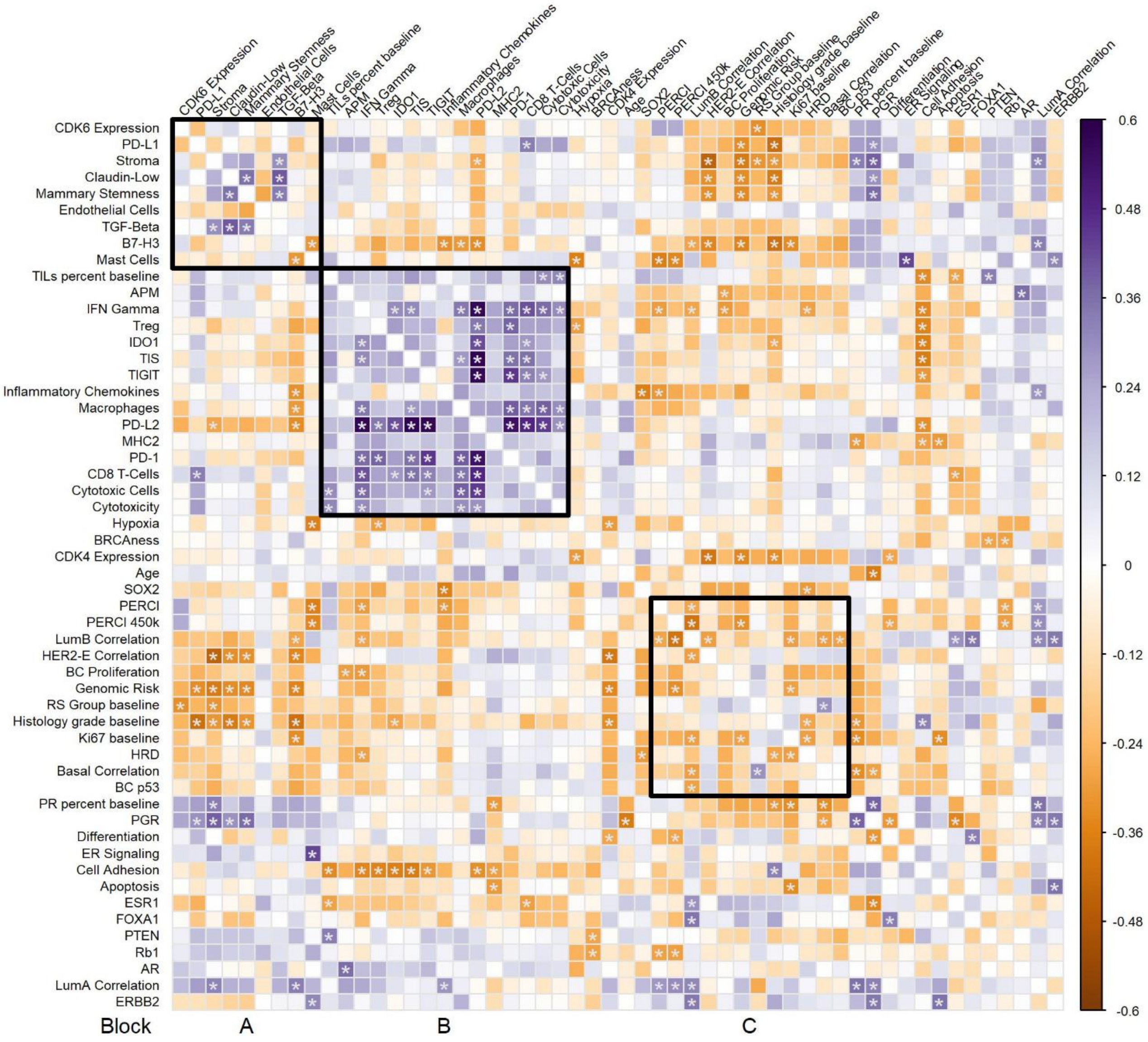


**Supplementary Fig. S7: Differences in signature score correlations in the NanoString TAM and AI groups**

Spearman's correlation coefficients between NanoString gene expression signatures and clinico-pathological parameters in the TAM and AI groups were compared using a Fisher's z-test. Effect sizes are shown as Cohen's q-values in a color range of orange (TAM > AI) to purple (AI > TAM). q-values of |0.10|, |0.30| and |0.50| are considered small, moderate, and large differences, respectively. Results with statistically significant differences between treatment groups are indicated by *, *p*-value < 0.05.

**Supplementary Fig. S8: Oncoprint of RGAs in the TCGA-BRCA subcohort**

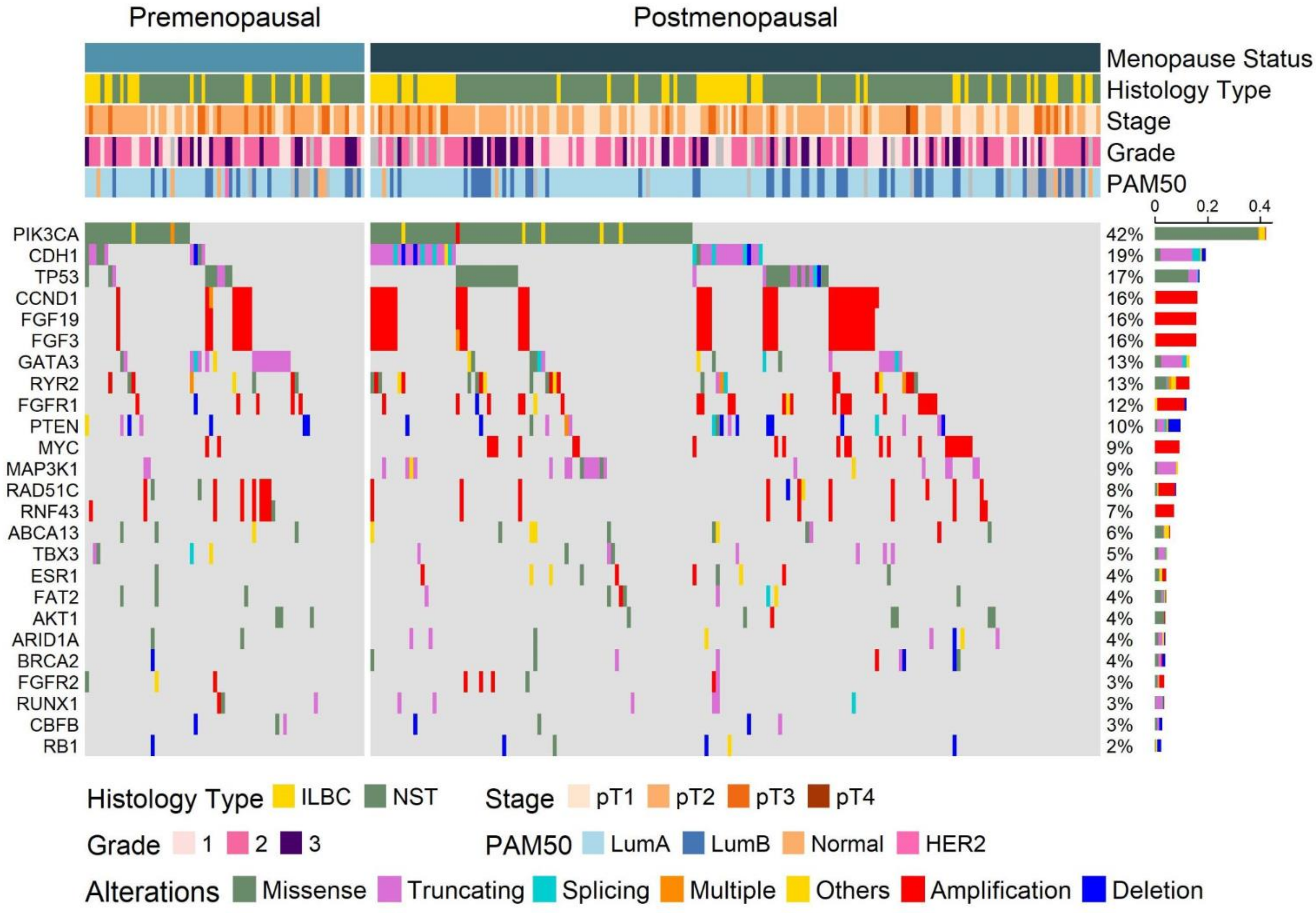


**Supplementary Fig. S8: Oncoprint of RGAs in the TCGA-BRCA subcohort**

The oncoprint summarizes the mutational landscape of the 25 most recurrent genomic alterations, derived from the NanoString cohort. The alterations are color-coded by RGA type and separated by menopause status. Clinical annotations of cases are indicated at the top of the figure. The bar plot on the right quantifies RGA recurrence, and genes are sorted by total alteration burden.

**Supplementary Fig. S9: RGAs resulting in altered gene expression in the NanoString cohort.**

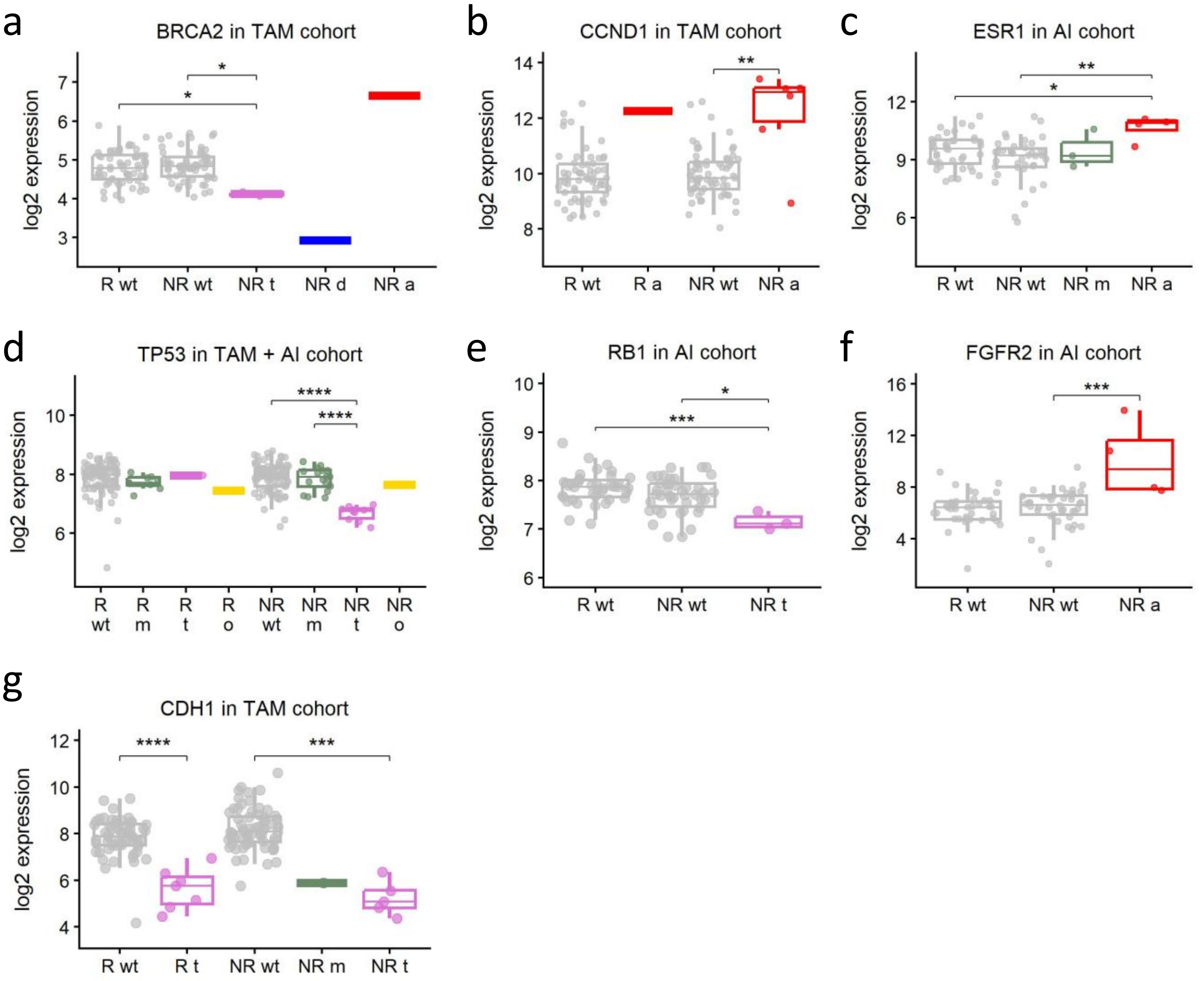


**Supplementary Fig. S9: RGAs resulting in altered gene expression in the NanoString cohort.** (a-g) NanoString log2 mRNA expression levels <u>at baseline before pET</u> of RGA with significant differences between NR and R groups, separated by RGA types and analyzed using Wilcoxon rank sum test with •, *, **, ***, ****, *p*-values < 0.1, 0.05, 0.01, 0.001, 0.0001. Expression data for CBFB and RYR2 are not available. Boxplots show median (line), upper, and lower quartiles (boxes), and lines extending to 1.5-IQR (whiskers). RGA types: amplification (a), deletion (d), missense (m), truncating (t), others (o).

**Supplementary Fig. S10: Differential expression of NanoString signatures between R and NR per RS risk groups.**

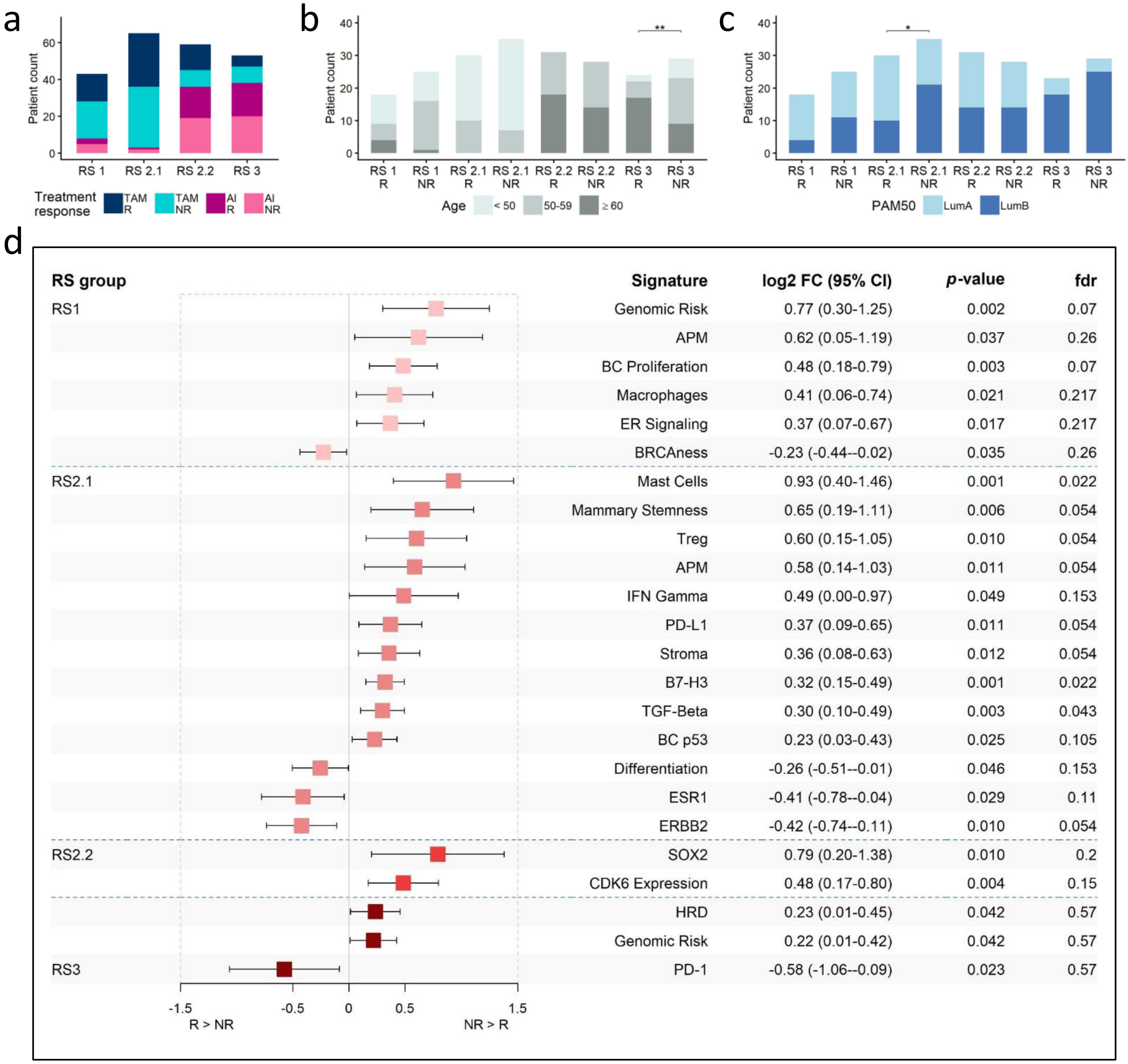


| RS group | Signature | log2 FC (95% CI) | *p*-value | fdr |
|---|---|---|---|---|
| RS1 | Genomic Risk | 0.77 (0.30-1.25) | 0.002 | 0.07 |
| | APM | 0.62 (0.05-1.19) | 0.037 | 0.26 |
| | BC Proliferation | 0.48 (0.18-0.79) | 0.003 | 0.07 |
| | Macrophages | 0.41 (0.06-0.74) | 0.021 | 0.217 |
| | ER Signaling | 0.37 (0.07-0.67) | 0.017 | 0.217 |
| | BRCAness | -0.23 (-0.44--0.02) | 0.035 | 0.26 |
| RS2.1 | Mast Cells | 0.93 (0.40-1.46) | 0.001 | 0.022 |
| | Mammary Stemness | 0.65 (0.19-1.11) | 0.006 | 0.054 |
| | Treg | 0.60 (0.15-1.05) | 0.010 | 0.054 |
| | APM | 0.58 (0.14-1.03) | 0.011 | 0.054 |
| | IFN Gamma | 0.49 (0.00-0.97) | 0.049 | 0.153 |
| | PD-L1 | 0.37 (0.09-0.65) | 0.011 | 0.054 |
| | Stroma | 0.36 (0.08-0.63) | 0.012 | 0.054 |
| | B7-H3 | 0.32 (0.15-0.49) | 0.001 | 0.022 |
| | TGF-Beta | 0.30 (0.10-0.49) | 0.003 | 0.043 |
| | BC p53 | 0.23 (0.03-0.43) | 0.025 | 0.105 |
| | Differentiation | -0.26 (-0.51--0.01) | 0.046 | 0.153 |
| | ESR1 | -0.41 (-0.78--0.04) | 0.029 | 0.11 |
| | ERBB2 | -0.42 (-0.74--0.11) | 0.010 | 0.054 |
| RS2.2 | SOX2 | 0.79 (0.20-1.38) | 0.010 | 0.2 |
| | CDK6 Expression | 0.48 (0.17-0.80) | 0.004 | 0.15 |
| | HRD | 0.23 (0.01-0.45) | 0.042 | 0.57 |
| | Genomic Risk | 0.22 (0.01-0.42) | 0.042 | 0.57 |
| RS3 | PD-1 | -0.58 (-1.06--0.09) | 0.023 | 0.57 |

**Supplementary Fig. S10: Differential expression of NanoString signatures between R and NR per RS risk groups.**

Stacked barplots indicating the distribution of treatment response (a), age groups (b), and PAM50 subtypes (c) per RS response groups. HER2-E sample is excluded in (c). Comparisons between Responder and Non-Responder groups were performed using a Cumulative Link Mixed Model for ordinal variables and a Generalized Linear Mixed-Effects Model for binary variables with patient pair ID as a random effect where applicable. Asterisks *, ** indicates *p*-values < 0.05, 0.01. (d) Differential signature expression between response groups per RS risk groups by fitting linear mixed models with pair-ids as random effect. The following variables are used as covariates: RS1: age; RS2.1: TILs group and PAM50 subtype; RS3: age. Only significant signatures with *p*-values < 0.05 are shown. Signatures are sorted by log2 fold change of mean signature expression between responders and non-responders per RS group. 95% confidence intervals of log2 fold change, *p*-values and fdr-adjusted *p*-values are shown.

**Supplementary Fig. S11: Associations between signatures and disease-free survival in the TAM group.**

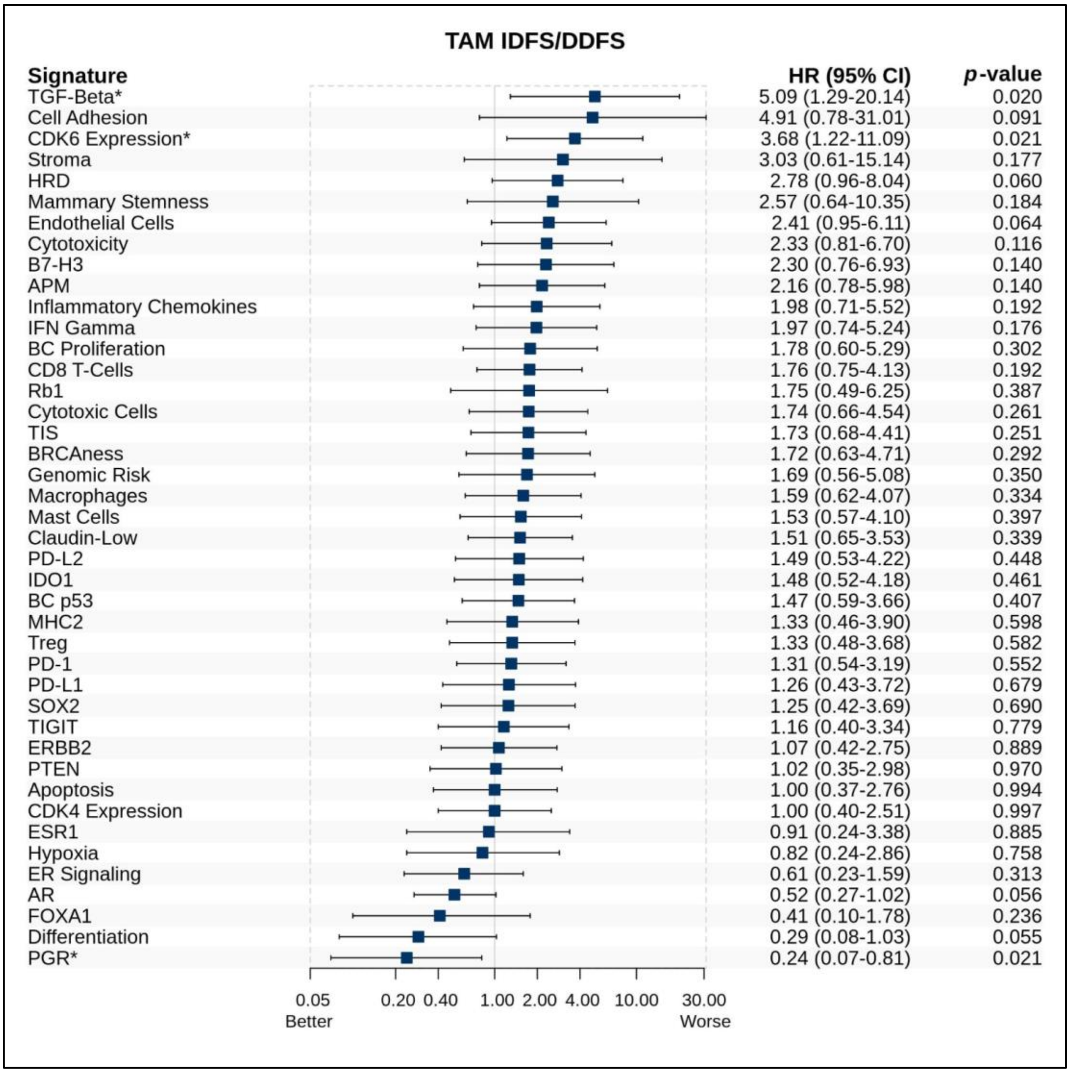


**Supplementary Fig. S11: Associations between BC360 signatures and disease-free survival in the TAM group.** Association of BC360 signatures with survival endpoints invasive and distant disease-free survival (IDFS/DDFS) in the TAM group were estimated by Cox regression and expressed as hazard ratios (HR) with 95% confidence intervals (95% CI) and *p*-values. N cases = 125, N events IDFS/DDFS = 4. Median follow up 59.8 months.
