## Supplementary Tables for "Integrative analysis of pre-treatment RNA expression signatures and recurrent genomic alterations: Link to menopausal status, short-term endocrine therapy response and disease-free survival in luminal breast cancer"

***Corresponding author:**

### **Table of contents:**

**Supplementary Table S1.** Clinico-pathologic characteristics of the NanoString cohort

**Supplementary Table S2.** Description of cohorts, data and results (xlsx file)

**Supplementary Table S3.** Clinico-pathologic features of the TCGA-BRCA sub-cohort

**Supplementary Table S4.** Clinico-pathologic comparison of the NanoString cohort and TCGA-BRCA sub-cohort

**Supplementary Table S5.** Summary of top 25 RGAs in the NanoString cohort

**Supplementary Table S6.** Summary of top 25 NanoString RGAs in the TCGA-BRCA subcohort

**Supplementary Table S7.** Univariate Cox regression analysis of mean signature expression and disease-free survival from KM-plotter

### **Supplementary Table S1. Clinico-pathologic characteristics of the NanoString cohort**

|  | **All**  **n=220** | **TAM R^A^**  **n=64** | **TAM NR^B^**  **n=71** | **AI R^A^**  **n=39** | **AI NR^B^**  **n=46** | ***p_TAM_*** | ***p_AI_*** | ***p_TAM-AI_*** |
| --- | --- | --- | --- | --- | --- | --- | --- | --- |
|  | n (%) | n (%) | n (%) | n (%) | n (%) |  |  |  |
| **Age** |  |  |  |  |  | *0.176* | ***0.004*** | ***3.4e-29*** |
| < 50 | 74 (34%) | 31 (46%) | 41 (58%) | 0 | 2 (4%) |  |  |  |
| 50 - 59 | 83 (37%) | 28 (44%) | 29 (40%) | 5 (12%) | 21 (46%) |  |  |  |
| ≥ 60 | 63 (29%) | 5 (10%) | 1 (2%) | 34 (88%) | 23 (50%) |  |  |  |
| **Histology, baseline** |  |  |  |  |  | *0.944* | *0.931* | ***0.046*** |
| NST | 201 (91%) | 57 (89%) | 62 (87%) | 38 (97%) | 44 (96%) |  |  |  |
| ILBC | 19 (9%) | 7 (11%) | 9 (13%) | 1 (3%) | 2 (4%) |  |  |  |
| **pT Stage** |  |  |  |  |  | *0.927***^C^** | *0.929***^C^** | *1***^C^** |
| pT1 | 141 (64%) | 42 (66%) | 44 (62%) | 24 (62%) | 31 (67%) |  |  |  |
| pT2 | 78 (35%) | 22 (34%) | 26 (37%) | 15 (38%) | 15 (33%) |  |  |  |
| pT3 | 1 (1%) | 0 | 1 (1%) | 0 | 0 |  |  |  |
| pT4 | 0 | 0 | 0 | 0 | 0 |  |  |  |
| **pN Stage** |  |  |  |  |  | *0.988* | *0.919* | *0.163* |
| pN0 | 206 (94%) | 61 (95%) | 68 (96%) | 36 (92%) | 41 (89%) |  |  |  |
| pN1+ | 14 (6%) | 3 (5%) | 3 (4%) | 3 (8%) | 5 (11%) |  |  |  |
| **Grade, baseline** |  |  |  |  |  | *0.943* | *0.899*^D^ | ***1.1e-08*** |
| G1 | 11 (5%) | 6 (9%) | 4 (6%) | 0 | 1 (2%) |  |  |  |
| G2 | 113 (51%) | 40 (63%) | 47 (66%) | 11 (28%) | 15 (33%) |  |  |  |
| G3 | 96 (44%) | 18 (28%) | 20 (28%) | 28 (72%) | 30 (65%) |  |  |  |
| **ER, baseline** |  |  |  |  |  | *1* | *1* | *1* |
| > 10% | 218 (99%) | 64 (100%) | 71 (100%) | 38 (97%) | 45 (98%) |  |  |  |
| ≤ 10% | 0 | 0 | 0 | 0 | 0 |  |  |  |
| n.a. | 2 (1%) | 0 | 0 | 1 (3%) | 1 (2%) |  |  |  |
| **PR, baseline** |  |  |  |  |  | *0.388* | *0.202* | ***5.2e-05*** |
| > 10% | 188 (85%) | 61 (95%) | 65 (92%) | 31 (79%) | 31 (67%) |  |  |  |
| ≤ 10% | 32 (15%) | 3 (5%) | 6 (8%) | 8 (21%) | 15 (33%) |  |  |  |
| **HER2 Status, baseline** |  |  |  |  |  | *1* | *0.459* | *1* |
| Negative | 218 (99%) | 64 (100%) | 70 (99%) | 38 (97%) | 46 (100%) |  |  |  |
| Positive | 2 (1%) | 0 | 1 (1%) | 1 (3%) | 0 |  |  |  |
| **E-cadherin, baseline** |  |  |  |  |  | *0.977* | *0.097* | ***0.039*** |
| pos. | 197 (89%) | 55 (86%) | 61 (86%) | 37 (95%) | 44 (96%) |  |  |  |
| neg. | 22 (10%) | 9 (14%) | 9 (13%) | 2 (5%) | 2 (4%) |  |  |  |
| n.a. | 1 (1%) | 0 | 1 (1%) | 0 | 0 |  |  |  |
| **Ki67, baseline** |  |  |  |  |  | *0.647* | *0.974* | ***6.6e-04*** |
| 0% - 9% | 14 (6%) | 7 (11%) | 5 (7%) | 1 (3%) | 1 (2%) |  |  |  |
| 10% - 19% | 53 (24%) | 18 (28%) | 21 (39%) | 7 (18%) | 7 (15%) |  |  |  |
| 20% - 34% | 127 (58%) | 34 (53%) | 39 (47%) | 23 (59%) | 31 (68%) |  |  |  |
| 35% - 100% | 26 (12%) | 5 (8%) | 6 (7%) | 8 (20%) | 7 (15%) |  |  |  |
| **Luminal Subtype^E^, baseline** | |  |  |  |  | *0.226* | *0.963* | ***7.3e-06*** |
| LumA | 164 (75%) | 57 (89%) | 58 (82%) | 22 (56%) | 27 (59%) |  |  |  |
| LumB | 56 (25%) | 7 (11%) | 13 (18%) | 17 (44%) | 19 (41%) |  |  |  |
| **PAM50^F^, baseline** |  |  |  |  |  | ***0.044***^C^ | *0.269*^C^ | ***0.002***^C^ |
| LumA | 102 (46%) | 41 (64%) | 33 (46%) | 15 (38%) | 13 (28%) |  |  |  |
| LumB | 117 (53%) | 23 (36%) | 38 (54%) | 23 (59%) | 33 (72%) |  |  |  |
| HER2-E | 1 (0%) | 0 | 0 | 1 (3%) | 0 |  |  |  |
| **TILs^G^, baseline** |  |  |  |  |  | ***0.031*^C^** | *0.580***^C^** | *0.452***^C^** |
| 0% - 9% | 160 (73%) | 43 (67%) | 59 (83%) | 25 (64%) | 33 (72%) |  |  |  |
| 10% - 40% | 53 (24%) | 20 (31%) | 11 (15%) | 11 (28%) | 11 (24%) |  |  |  |
| 41% - 100% | 3 (1%) | 1 (2%) | 0 | 0 | 2 (4%) |  |  |  |
| n.a. | 4 (2%) | 0 | 1 (2%) | 3 (8%) | 0 |  |  |  |
| **Oncotype DX RS Group, baseline** | |  |  |  |  | *0.941* | *0.949* | ***3.4e-08*** |
| 1 (0 -11) | 43 (20%) | 15 (23%) | 20 (28%) | 3 (8%) | 5 (11%) |  |  |  |
| 2 (12 – 25) | 124 (56%) | 43 (67%) | 42 (59%) | 18 (46%) | 21 (46%) |  |  |  |
| 3 (26 – 100) | 55 (24%) | 6 (10%) | 9 (13%) | 18 (46%) | 20 (43%) |  |  |  |

Unless otherwise stated, the values are given in the format n (%), with n corresponding to the number of patients. Comparisons between Responder and Non-Responder groups were performed using a Cumulative Link Mixed Model for ordinal variables and a Generalized Linear Mixed-Effects Model for binary variables with patient pair ID as a random effect where applicable. For sparse data with complete or near separation, Fisher’s exact test was applied. For the statistical analysis between TAM and AI groups, the chi-squared test for trends was used to compare Ki67 at baseline, and Fisher's exact test was used for all other comparisons. Significant differences are highlighted in bold. Cases with missing data were excluded from each statistical test.

n.a. not applicable

^A^Responder was defined as post-pET Ki67 < 10% and relative Ki67 decrease of ≤ -70% from baseline to post-pET.

^B^Non-reponder was defined as post-pET Ki67 of ≥ 20% and relative Ki67 decrease of > -20% from baseline to post-pET.

^C^Comparison between the first two category groups only.

^D^Comparison between the last two category groups only.

^E^Luminal B was defined as Ki67 baseline ≥ 35% or PR baseline ≤ 20%, all other cases are categorised as Luminal A.

^F^Intrinsic subtypes of breast cancer using PAM50 gene expression signatures.

^G^Stromal tumor-infiltrating lymphocytes in pathologic tissue sections.

### **Supplementary Table S3. Clinico-pathologic features of the TCGA-BRCA sub-cohort**

|  | **All**  **n=260** | **Premenopausal**  **n=72** | **Postmenopausal**  **n=188** | ***p_Pre-Post_^A^*** |
| --- | --- | --- | --- | --- |
|  | n (%) | n (%) | n (%) |  |
| **Age** |  |  |  | ***1.6e-39*** |
| < 50 | 69 (27%) | 59 (82%) | 10 (5%) |  |
| 50 - 59 | 75 (29%) | 13 (18%) | 62 (33%) |  |
| ≤ 60 | 116 (44%) | 0 | 116 (62%) |  |
| **Histology** |  |  |  | *0.654* |
| NST | 181 (70%) | 52 (72%) | 129 (69%) |  |
| ILBC | 79 (30%) | 20 (28%) | 59 (31%) |  |
| **pT Stage** |  |  |  | ***0.035***^B^ |
| pT1 | 76 (29%) | 13 (18%) | 63 (33%) |  |
| pT2 | 150 (58%) | 47 (65%) | 103 (55%) |  |
| pT3 | 33 (13%) | 12 (17%) | 21 (11%) |  |
| pT4 | 1 (0%) | 0 | 1 (1%) |  |
| **pN Stage** |  |  |  | *0.103* |
| pN0 | 123 (47%) | 28 (39%) | 95 (51%) |  |
| pN1+ | 137 (53%) | 44 (61%) | 93 (49%) |  |
| **Grade** |  |  |  | *0.444*^B^ |
| G1 | 74 (28%) | 18 (25%) | 56 (30%) |  |
| G2 | 121 (47%) | 37 (51%) | 84 (45%) |  |
| G3 | 50 (19%) | 15 (21%) | 35 (18%) |  |
| n.a. | 15 (6%) | 2 (3%) | 13 (7%) |  |
| **ER (IHC)** |  |  |  | *1* |
| Positive | 260 (100%) | 72 (100%) | 188 (100%) |  |
| Negative | 0 | 0 | 0 |  |
| **PR (IHC)** |  |  |  | *1*^C^ |
| Positive | 227 (87%) | 64 (89%) | 163 (87%) |  |
| Negative | 32 (12%) | 8 (11%) | 24 (13%) |  |
| Indeterminate | 1 (1%) | 0 | 1 (0%) |  |
| **HER2 (IHC)** |  |  |  | *1*^C^ |
| Negative | 186 (72%) | 52 (72%) | 134 (71%) |  |
| Equivocal^D^ | 74 (28%) | 20 (28%) | 54 (29%) |  |
| Positive | 0 | 0 | 0 |  |
| **PAM50 Subtype** |  |  |  | *0.732*^C^ |
| LumA | 169 (65%) | 45 (63%) | 124 (66%) |  |
| LumB | 59 (23%) | 14 (19%) | 45 (24%) |  |
| Normal | 9 (3%) | 5 (7%) | 4 (2%) |  |
| HER2 | 1 (1%) | 1 (1%) | 0 |  |
| n.a. | 22 (8%) | 7 (10%) | 15 (8%) |  |
| **Race** |  |  |  | ***0.032*** |
| White | 222 (86%) | 55 (76%) | 167 (89%) |  |
| Asian | 11 (4%) | 4 (6%) | 7 (4%) |  |
| Black^E^ | 27 (10%) | 13 (18%) | 14 (7%) |  |
| **PFS status^F^** |  |  |  | *0.080* |
| 0 | 239 (92%) | 63 (88%) | 176 (94%) |  |
| 1 | 21 (8%) | 9 (12%) | 12 (6%) |  |

Unless otherwise stated, the values are given in the format n (%), with n corresponding to the number of patients. For the statistical analysis between pre- and postmenopausal groups, the Fisher’s exact test (FET) was used. Significant differences are highlighted in bold.

N.a. not applicable, PFS progression-free survival.

^A^Comparison between premenopausal (Pre) and postmenopausal (Post) groups.

^B^Comparison between the first three category groups only.

^C^Comparison between the first two category groups only.

^D^Her2 equivocal was defined with scores of 2+ in the immunohistochemistry (IHC) assay or with a HER2/CEP17 ratio between 1.8 and 2.2 in Fluorescence in situ hybridization (FISH).

^E^Black or African American

^F^PFS status: 0 censored, 1 progression

### **Supplementary Table S4. Clinico-pathologic comparison of the NanoString cohort and TCGA-BRCA sub-cohort**

|  | **NanoString TAM**  **n=135** | **TCGA-BRCA  Pre**  **n=72** | **NanoString**  **AI**  **n=85** | **TCGA-BRCA**  **Post**  **n=188** | ***P_Pre_*** | ***P_Post_*** | ***P_NanoString-TCGA_*** |
| --- | --- | --- | --- | --- | --- | --- | --- |
|  | n (%) | n (%) | n (%) | n (%) |  |  |  |
| **Age** |  |  |  |  | ***7.1e-05*** | *0.508* | ***0.001*** |
| < 50 | 72 (53%) | 59 (82%) | 2 (2%) | 10 (5%) |  |  |  |
| 50 - 59 | 57 (42%) | 13 (18%) | 26 (31%) | 62 (33%) |  |  |  |
| ≥ 60 | 6 (4%) | 0 (0%) | 57 (67%) | 116 (62%) |  |  |  |
| **Histology, baseline** |  |  |  |  | ***0.006*** | ***3.9e-08*** | ***1.8e-09*** |
| NST | 119 (88%) | 52 (72%) | 82 (96%) | 129 (69%) |  |  |  |
| ILBC | 16 (12%) | 20 (28%) | 3 (4%) | 59 (31%) |  |  |  |
| **pT Stage** |  |  |  |  | ***5.1e-12*** | ***2.8e-07*** | ***1.8e-17*** |
| pT1 | 86 (64%) | 13 (18%) | 55 (65%) | 63 (34%) |  |  |  |
| pT2 | 48 (36%) | 47 (65%) | 30 (35%) | 103 (55%) |  |  |  |
| pT3 | 1 (1%) | 12 (17%) | 0 | 21 (11%) |  |  |  |
| pT4 | 0 | 0 | 0 | 1 (1%) |  |  |  |
| **pN Stage** |  |  |  |  | ***1.8e-12*** | ***2.5e-11*** | ***1.4e-30*** |
| pN0 | 129 (96%) | 28 (39%) | 77 (91%) | 95 (51%) |  |  |  |
| pN1+ | 6 (4%) | 44 (61%) | 8 (9%) | 93 (49%) |  |  |  |
| **Grade** |  |  |  |  | ***7.3e-04*** | ***1.2e-16*** | ***6.5e-15*** |
| G1 | 10 (7%) | 18 (25%) | 1 (1%) | 56 (30%) |  |  |  |
| G2 | 87 (64%) | 37 (51%) | 26 (31%) | 84 (45%) |  |  |  |
| G3 | 38 (28%) | 15 (21%) | 58 (68%) | 35 (19%) |  |  |  |
| n.a. | 0 | 2 (3%) | 0 | 13 (7%) |  |  |  |
| **ER Status** |  |  |  |  | *1* | *1* | *1* |
| Positive | 134 (99%) | 72 (100%) | 83 (98%) | 188 (100%) |  |  |  |
| Negative | 0 | 0 | 0 | 0 |  |  |  |
| n.a. | 1 (1%) | 0 | 2 (2%) | 0 |  |  |  |
| **PR Status** |  |  |  |  | ***0.018*** | *0.144* | *0.303* |
| Positive | 132 (98%) | 64 (89%) | 68 (80%) | 163 (87%) |  |  |  |
| Negative | 3 (2%) | 8 (11%) | 17 (20%) | 24 (13%) |  |  |  |
| Indeterminate | 0 | 0 | 0 | 1 (0%) |  |  |  |
| **HER2 Status** |  |  |  |  | ***9.4e-11*** | ***2.8e-10*** | ***1.1e-22*** |
| Negative | 134 (99%) | 52 (72%) | 84 (99%) | 134 (71%) |  |  |  |
| Equivocal | 0 | 20 (28%) | 0 | 54 (29%) |  |  |  |
| Positive | 1 (1%) | 0 | 1 (1%) | 0 |  |  |  |
| **PAM50, baseline** |  |  |  |  | ***1.3e-07*** | ***1.2e-09*** | ***2.8e-09*** |
| LumA | 74 (55%) | 45 (62%) | 28 (33%) | 124 (66%) |  |  |  |
| LumB | 61 (45%) | 14 (19%) | 56 (66%) | 45 (24%) |  |  |  |
| Normal | 0 | 5 (7%) | 0 | 4 (2%) |  |  |  |
| HER2 | 0 | 1 (1%) | 1 (1%) | 0 |  |  |  |
| n.a. | 0 | 7 (10%) | 0 | 15 (8%) |  |  |  |

Unless otherwise stated, the values are given in the format n (%), with n corresponding to the number of patients. For the statistical analysis between NanoString and TCGA sub-cohort groups, Fisher's exact test was used. Significant differences are highlighted in bold.

### **Supplementary Table S5.** **Summary of top 25 RGAs in the NanoString cohort**

| **Gene** | **All**  **n=220** | **TAM**  **n=135** | **AI**  **n=85** | **p-value** | **fdr** |
| --- | --- | --- | --- | --- | --- |
|  | n (%) | n (%) | n (%) |  |  |
| **PIK3CA** | 98 (44.5) | 63 (46.7) | 35 (41.2) | *0.255* | *0.861* |
| **GATA3^A^** | 49 (22.3)** | 35 (25.9) | 14 (16.5) | *0.069* | *0.776* |
| **TP53** | 39 (17.7) | 19 (14.1) | 20 (23.5) | *0.055* | *0.776* |
| **MAP3K1** | 34 (15.5)* | 29 (21.5)** | 5 (5.9) | ***0.001*** | *0.064* |
| **CBFB** | 26 (11.8)** | 20 (14.8)* | 6 (7.1) | *0.061* | *0.776* |
| **FGF19** | 21 (9.5) | 8 (5.9) | 13 (15.3) | ***0.021*** | *0.776* |
| **FGF3** | 20 (9.1) | 9 (6.7) | 11 (12.9) | *0.092* | *0.822* |
| **ABCA13** | 18 (8.2) | 9 (6.7) | 9 (10.6) | *0.216* | *0.861* |
| **FGFR1** | 18 (8.2) | 10 (7.4) | 8 (9.4) | *0.386* | *0.861* |
| **RYR2** | 17 (7.7) | 8 (5.9) | 9 (10.6) | *0.158* | *0.834* |
| **CCND1** | 16 (7.3) | 7 (5.2) | 9 (10.6) | *0.109* | *0.834* |
| **CDH1** | 16 (7.3) | 13 (9.6) | 3 (3.5) | *0.073* | *0.776* |
| **FAT2** | 16 (7.3) | 7 (5.2) | 9 (10.6)* | *0.109* | *0.834* |
| **AKT1** | 13 (5.9) | 9 (6.7) | 4 (4.7) | *0.397* | *0.861* |
| **PTEN** | 12 (5.5) | 5 (3.7) | 7 (8.2) | *0.129* | *0.834* |
| **MYC** | 11 (5.0) | 6 (4.4) | 5 (5.9) | *0.429* | *0.861* |
| **TBX3** | 11 (5.0) | 5 (3.7) | 6 (7.1) | *0.212* | *0.861* |
| **BRCA2** | 10 (4.5) | 6 (4.4) | 4 (4.7) | *0.586* | *0.861* |
| **RUNX1** | 10 (4.5) | 4 (3.0) | 6 (7.1) | *0.139* | *0.834* |
| **ARID1A** | 9 (4.1) | 6 (4.4) | 3 (3.5) | *0.516* | *0.861* |
| **ESR1** | 7 (3.2) | 0 (0) | 7 (8.2) | ***0.001*** | *0.064* |
| **RNF43** | 7 (3.2) | 2 (1.5) | 5 (5.9) | *0.080* | *0.776* |
| **RAD51C** | 6 (2.7) | 1 (0.7) | 5 (5.9) | ***0.033*** | *0.776* |
| **RB1** | 6 (2.7) | 2 (1.5) | 4 (4.7) | *0.158* | *0.834* |
| **FGFR2** | 5 (2.3) | 1 (0.7) | 4 (4.7) | *0.077* | *0.776* |

Counts and frequencies of top 25 RGA in the NanoString cohort. Significant differences between treatment groups were analyzed using Fisher's exact test and *p*-values and fdr-adjusted *p*-values are shown. Significant differences between TAM and AI are highlighted in bold print. For All, frequencies of the TAM and AI cohorts were combined.

^A^*GATA3* splicing mutations were significantly enriched in the TAM cohort (TAM: n=17 (12.6%), AI: n=1 (1.2%), *p*-value=*0.001*)

**: significantly more frequent than in the same group of the TCGA-BRCA subcohort, fdr<0.05

*: significantly more frequent than in the same group of the TCGA-BRCA subcohort, *p*-value<0.05

### **Supplementary Table S6.** **Summary of top 25 NanoString RGAs in the TCGA-BRCA subcohort**

| **Gene** | **All**  **n=260** | **Premenopausal**  **n=72** | **Postmenopausal**  **n=188** | **p-value** | **fdr** |
| --- | --- | --- | --- | --- | --- |
|  | n (%) | n (%) | n (%) |  |  |
| **PIK3CA** | 110 (42.5) | 27 (37.5) | 83 (44.4) | *0.194* | *0.373* |
| **CDH1** | 50 (19.2)** | 10 (13.9) | 40 (21.3)** | *0.118* | *0.350* |
| **TP53** | 44 (17.0) | 10 (13.9) | 34 (18.2) | *0.265* | *0.473* |
| **CCND1** | 42 (16.2)** | 8 (11.1) | 34 (18.2) | *0.114* | *0.350* |
| **FGF19** | 41 (15.8)* | 8 (11.1) | 33 (17.6) | *0.134* | *0.350* |
| **FGF3** | 41 (15.8)* | 8 (11.1) | 33 (17.6) | *0.134* | *0.350* |
| **GATA3** | 34 (13.1) | 17 (23.6) | 17 (9.1) | ***0.003*** | *0.067* |
| **RYR2** | 34 (13.1)* | 8 (11.1) | 26 (13.9) | *0.355* | *0.555* |
| **FGFR1** | 31 (12.0) | 6 (8.3) | 25 (13.4) | *0.184* | *0.373* |
| **PTEN** | 25 (9.7) | 7 (9.7%) | 18 (9.6) | *0.573* | *0.617* |
| **MYC** | 24 (9.3) | 2 (2.8) | 22 (11.8) | ***0.016*** | *0.181* |
| **MAP3K1** | 23 (8.9) | 2 (2.8) | 21 (11.2) | ***0.022*** | *0.181* |
| **RAD51C** | 21 (8.1)** | 9 (12.5)** | 12 (6.4) | *0.091* | *0.350* |
| **RNF43** | 19 (7.3)* | 9 (12.5)** | 10 (5.3) | ***0.048*** | *0.299* |
| **ABCA13** | 15 (5.8) | 4 (5.6) | 11 (5.9) | *0.592* | *0.617* |
| **TBX3** | 12 (4.6) | 4 (5.6) | 8 (4.3) | *0.439* | *0.570* |
| **ESR1** | 11 (4.2) | 1 (1.4) | 10 (5.3) | *0.140* | *0.350* |
| **FAT2** | 11 (4.2) | 3 (4.2) | 8 (4.3) | *0.632* | *0.632* |
| **AKT1** | 10 (3.9) | 3 (4.2) | 7 (3.7) | *0.559* | *0.617* |
| **ARID1A** | 10 (3.9) | 2 (2.8) | 8 (4.3) | *0.441* | *0.570* |
| **BRCA2** | 10 (3.9) | 1 (1.4) | 9 (4.8) | *0.181* | *0.373* |
| **FGFR2** | 9 (3.5) | 3 (4.2) | 6 (3.2) | *0.479* | *0.570* |
| **RUNX1** | 9 (3.5) | 3 (4.1) | 6 (3.2) | *0.479* | *0.570* |
| **CBFB** | 7 (2.7) | 3 (4.2) | 4 (2.1) | *0.300* | *0.500* |
| **RB1** | 6 (2.3) | 1 (1.4) | 5 (2.7) | *0.467* | *0.570* |

Counts and frequencies of the top 25 NanoString RGAs in the TCGA-BRCA subcohort. Significant differences between menopausal groups were analyzed using Fisher-exact test and *p*-values and fdr-adjusted *p*-values are shown. Significant differences between menopause groups are highlighted in bold print. For All, frequencies of the pre- and postmenopausal groups were combined.

**: significantly more frequent than in the same group of the NanoString cohort, fdr<0.05

*: significantly more frequent than in the same group of the NanoString cohort, *p*-value<0.05

### **Supplementary Table S7. Univariate Cox regression analysis of mean signature expression and disease-free survival from KM-plotter**

|  | **Signature** | **HR (95% CI)^A^** | ***p*-value** | **n** |
| --- | --- | --- | --- | --- |
| **RFS^B^** | |  |  |  |
|  | Hypoxia* | 2.17 (1.45 - 3.25) | ***0.0002*** | 764 |
|  | HRD* | 1.53 (1.07 - 2.18) | ***0.0187*** | 764 |
|  | BC p53* | 1.47 (1.03 - 2.10) | ***0.0339*** | 764 |
|  | PD-1 | 1.03 (0.84 - 1.25) | *0.7943* | 2301 |
|  | ERBB2* | 0.77 (0.62 - 0.94) | ***0.0127*** | 2301 |
|  | PGR* | 0.52 (0.36 - 0.75) | ***0.0004*** | 2301 |
| **DMFS** |  |  |  |  |
|  | BC p53* | 3.19 (1.12 - 9.10) | ***0.0301*** | 183 |
|  | Hypoxia | 2.33 (0.73 - 7.42) | *0.1537* | 183 |
|  | PD-1 | 1.28 (0.89 - 1.83) | *0.1828* | 1002 |
|  | ERBB2* | 0.69 (0.48 - 1.00) | ***0.0475*** | 1002 |
|  | HRD | 0.49 (0.17 - 1.42) | *0.1899* | 183 |
|  | PGR* | 0.25 (0.08 - 0.79) | ***0.0182*** | 1002 |
| **OS** |  |  |  |  |
|  | HRD | 7.09 (0.94 - 53.64) | *0.0577* | 182 |
|  | Hypoxia | 2.01 (0.74 - 5.50) | *0.1725* | 182 |
|  | ERBB2 | 0.73 (0.46 - 1.15) | *0.1764* | 630 |
|  | BC p53 | 0.62 (0.22 - 1.75) | *0.3707* | 182 |
|  | PD-1* | 0.55 (0.34 - 0.89) | ***0.0138*** | 630 |
|  | PGR* | 0.17 (0.04 - 0.77) | ***0.0215*** | 630 |

Signatures with significant associations with survival in the NanoString AI group are shown (except for Genomic Risk, which does not represent a mean expression value). PGR might be more relevant in premenopausal cases with significantly higher expression of PGR. Data from KM-plotter^2^ Breast cancer (mRNA, gene chip, last accessed on 2025-06-25) was filtered using the following criteria: mean of expression of all genes per signature; auto select best cut-off; 60 months follow-up time; only JetSet best probe set; default settings for quality control. The analysis was restricted to ER+/HER2- cases which had received endocrine therapy and/or chemotherapy. mRNA expression of MKI67 and ESR1 was used as covariate. Results are not corrected for multiple testing. Asterisks and bold print indicate significant results with *p*-value < 0.05.

^A^Hazard ratio (95% confidence interval, lower limit, upper limit).

^B^RFS: recurrence-free survival, DMFS: distant metastasis-free survival, OS: overall survival
